# Mucosal and Systemic Antibodies Associated with Clinical Protection in a Pertussis Controlled Human Infection Model

**DOI:** 10.64898/2026.06.12.26355530

**Authors:** Patricia L. Milletich, May ElSherif, Kirsten Marx, Amanda Caulfield, Susan Hariri, Scott Halperin, Marcela F. Pasetti

**Author notes:** Corresponding Author: Marcela F. Pasetti. **Disclaimer**: The findings and conclusions in this report are those of the authors and do not necessarily represent the official position of the Centers for Disease Control and Prevention, US Department of Health and Human Services.

## Abstract

**Background:** The engagement of mucosal and systemic immunity in preventing *Bordetella pertussis* colonization and infection in humans, the impact of prior vaccination on host immunity and protective outcomes, and the dynamics of the host response following exposure remain poorly understood.

**Methods:** Healthy adults were challenged with increasing colony-forming units (CFUs) doses, 10^6^-10^8^, of *B. pertussis* D420 intranasally (NCT05136599). Shedding (PCR and culturing) and symptom development were monitored up to 21 days post-challenge. Serum and nasal wash IgA and IgG were measured before challenge (baseline) and up to 6 months post-challenge.

**Findings:** Antibodies increased post-challenge only in infected individuals, primarily nasal IgA. Participants who remained uninfected had higher baseline levels of filamentous hemagglutinin (FHA)- specific mucosal IgA and IgG, and higher serum IgA against fimbriae 2/3 (FIM). FHA was negatively associated with bacterial load and was a key discriminator between shedders and non-shedders, up to one week post-challenge. By day 14 post-challenge, pertussis toxin (PT) IgG and FIM IgA in both serum and mucosal samples were negatively associated with bacterial colonization. The majority (96·7%) of acellular pertussis (aP) vaccine recipients (n=23, median age 22·0 years) became infected, compared to 69·4% of those who received whole-cell pertussis vaccine (n=36; median age 32·0 years), and their antibody responses remained distinct following infection.

**Interpretation:** Nasal FHA antibodies emerged as early predictors of protection against pertussis infection, while PT IgG and FIM IgA antibodies may reflect clearance after infection. aP-primed individuals were more susceptible to infection, despite their younger age and more recent vaccination.

**Funding:** CDC Contract #75D30122C15467 and CDC IPA Agreement #24IPA2417512

**Research in Context:** *Evidence before this study:* Immunological correlates of protection against pertussis infection remain elusive. The contributions of mucosal and systemic immunity in preventing infection in humans, the impact of prior vaccination on risk of infection and clinical outcomes, and the recall response following exposure remain incompletely understood. We retrieved PubMed literature in any language since inception through February 1st, 2026, using the terms: (“Pertussis” OR “Whooping Cough”) AND (“IgA” or “immunoglobulin” OR “IgG”) AND (“Coloni*ation” OR “adherence” OR “Clearance” OR “shed*” OR “Challenge”) AND “Human”. A total of 44 publications were identified, two of which report data from controlled human infection model (CHIM) studies. The PERISCOPE CHIM report examined only the serum *B. pertussis* antibody responses after infection, and the other evaluated the efficacy of the live-attenuated vaccine BPZE1 in preventing *B. pertussis* colonization. Neither of these studies examined the dynamics of mucosal and systemic antibodies before and after challenge in relation to bacterial colonization and symptomatic illness, nor did they investigate the influence of prior childhood vaccine priming (acellular pertussis [aP] vs. whole-cell pertussis [wP]) on clinical outcomes. The host immune factors that determine susceptibility to pertussis infection remain unknown.

*Added Value of this study:* We report here that baseline mucosal and systemic antibodies specific to filamentous hemagglutinin (FHA) were associated with clinical protection against pertussis infection among CHIM participants. Two weeks after infection, mucosal and serum pertussis toxin (PT) IgG and Fimbriae 2/3 IgA were negatively correlated with pertussis colonization. Primary vaccine history (whole-cell [wP] or acellular [aP]) imprinted immune profiles that persisted after experimental infection in adulthood and affected clinical outcomes. This study is the first to identify mucosal and systemic temporal markers of pertussis infection and predictors of protective immunity in humans.

*Implications of all available evidence:* The insights generated by these results on host immunity associated with protection against pertussis, particularly the contribution of mucosal FHA antibodies, can inform vaccine design and guide population assessments of susceptibility to infection.

## INTRODUCTION

*Bordetella pertussis* causes a respiratory disease (known as pertussis, or whooping cough) that is prevalent worldwide and disproportionately affects young children. Whole-cell pertussis (wP) vaccines were introduced in the 1940s, and the acellular pertussis (aP) vaccine became available in the 1990s. Despite high global vaccination coverage, some countries are experiencing a resurgence of pertussis, as evidenced by the increase in cases in the United States from 2023 to 2024.^2^ It is theorized that asymptomatic infections and waning immunity contribute to the reappearance of pertussis infection in adolescents and adults.^3,4^

The coordinated features of *B. pertussis* immunity that prevent symptomatic upper respiratory tract infection, and the specific contributions of mucosal and systemic antibodies, remain unclear. Given evidence of waning immunity in aP-vaccinated individuals, the population’s susceptibility to pertussis infection is a public health concern.^1^ In a pertussis baboon model, mucosal antibodies were associated with reduced or no colonization.^5^ There is a paucity of information on mucosal immunity to pertussis infection in humans, particularly regarding the target antigens and immunoglobulin classes that can exert distinct antimicrobial functions and interrupt bacterial colonization and illness.

Controlled human infection models (CHIMs) offer a powerful approach for studying host–pathogen interactions under highly regulated conditions. Two CHIM studies have been conducted in Europe in recent years. The CHIM study led by Read et al. (PERItussiS Correlates Of Protection Europe - Periscope) examined systemic antibodies only in relation to *B. pertussis* colonization – not symptomatic infection.^6,7^ The other CHIM study challenged participants who had been vaccinated with the live-attenuated pertussis vaccine (BPZE1) to evaluate its immunogenicity and efficacy – its purpose was not to dissect principles of infection and immunity.^8^

In this manuscript, we report results from a comprehensive analysis of mucosal and systemic antibodies in healthy adults enrolled in a pertussis CHIM study at the Canadian Center for Vaccinology, Dalhousie University (Halifax, Nova Scotia, Canada).^9^ Expanding on previous CHIM studies, the Dalhousie study involved participants with different vaccination histories and higher challenge doses and evaluated pertussis infection with and without symptoms. Herein, we investigated the dynamics of serum IgG and nasal wash IgG and IgA in individuals challenged with increasing doses of *B. pertussis* and monitored up to 6 months post-challenge, and their association with bacterial shedding and symptomatic illness. We also examined the impact of vaccination history (aP vs wP) on immune responses and challenge outcomes.

## METHODS

### Participants

In the initial study (NCT05136599), healthy adults with PT serum IgG<20 ELISA units (EU)/mL were enrolled. 75 participants received a single intranasal *B. pertussis* 420 inoculation at seven doses: 10^4^, 10^5^, 5×10^5^, 10^6^, 5×10^6^, 10^7^, 5×10^7^, and 10^8^ colony-forming units (CFU).^10^ Serum and nasal wash samples were collected on days -1, 1, 3, 7, 14, 28, and 56. Additionally, serum samples were collected on days 84 and 180 post-challenge. Participants who received inoculation doses of 10^4^, 10^5^, or 5×10^5^ CFU were excluded from the current antibody analysis due to low bacterial detection rates (0%, 40%, and 20%, respectively).

### Procedures

Shedding was determined by PCR positivity and bacterial colonization (CFU) in nasal washes during the inpatient observation period (through 16-21 days post-challenge).

Full details of the immunological assays are provided in the **Appendix pp. 3-5**. Briefly, serum IgG and nasal wash IgA and IgG against PT, PRN, FHA, and FIM were quantified using a MesoScale Discovery (MSD) multiplex assay that reports results in International Units (IU)/mL.^20^ ACT was run in a separate ELISA and reported in EU/mL. For mucosal samples, total IgA and IgG antibody levels were measured, and the titers were normalized (IU/µg or EU/µg) before analysis.

### Outcomes

Participants were classified as positive for *B. pertussis* shedding if they had ≥1 positive culture and/or ≥3 positive PCRs on or after day six post-challenge.^10^ Symptomatic participants were positive for pertussis and had ≥2 symptoms, including ≥1 respiratory symptom. Asymptomatic participants were positive for pertussis but had no respiratory symptoms. Non-infected participants tested negative for pertussis.

Seroconversion and positive responses in serum and nasal samples, respectively, were defined as a fold-change >4 relative to baseline.

### Statistical Analysis

Statistical analysis was performed using RStudio (version 4.5.2). R packages used for the analysis are listed in **Appendix pp. 5**.

When comparing serum samples across dose groups, samples from the 5×10^6^ cohort were filtered to exclude days with fewer than half of the participants present (days 1, 3, 6, and 84 post-challenge), and two participants were additionally excluded due to a low number of time points. Pearson correlations were computed between each participant’s peak titers and the inoculation dose.

Fold-change was calculated relative to the baseline for all available samples. Only participants with a sample collected more than 14 days post-challenge were included in the percentages of participants with a positive response (≥4-fold change) across variables, which were compared using Fisher’s exact test.

Baseline titers of antigens – PT, PRN, FIM, and FHA – were compared through the Friedman test, and the comparison of PT to other antigens was performed through the paired Wilcoxon test. ACT was excluded due to differences in assay units (IU vs EU). PLS-DA analysis was performed on baseline titers to compare clinical outcomes or vaccine priming.

Associations between colony counts from nasal washes and antibody titers were assessed using Kendall’s tau rank correlation, and the correlation between serum IgG and nasal IgG was assessed through Pearson correlation.

Thresholds for discriminatory performance were defined as AUC≥0·70, Youden index≥0·30, sensitivity>0·70, and specificity>0·50.

## RESULTS

75 participants enrolled in the CHIM study were challenged with 1×10^4^ to 1×10^8^ colony-forming units (CFU) of *B. pertussis* D420 (NCT05136599) in a dose-escalation approach.^10^ Groups that received the lowest dosage levels (10^4^, 10^5^, and 5×10^5^ CFU, n=16) were excluded from analysis due to low infection rates (0%, 40%, and 20%, respectively). The remaining 59 individuals received increasing doses of *B. pertussis* D420: 10^6^ CFU (n=6), 5×10^6^ CFU (n=10), 10^7^ CFU (n=22), 5×10^7^ CFU (n=12), and 10^8^ CFU (n=9). Mucosal and blood samples were collected longitudinally up to 56 days and 6 months post-challenge, respectively, and tested for antibodies against pertussis antigens – pertussis toxin (PT), pertactin (PRN), filamentous hemagglutinin (FHA), fimbriae 2/3 (FIM), and adenylate cyclase toxin (ACT) (**figure 1A)**. A quantitative analysis of serum and mucosal IgA and IgG was conducted using qualified Meso Scale Discovery (MSD) multiplex assays and enzyme-linked immunosorbent assays (ELISA).

**Figure 1.**
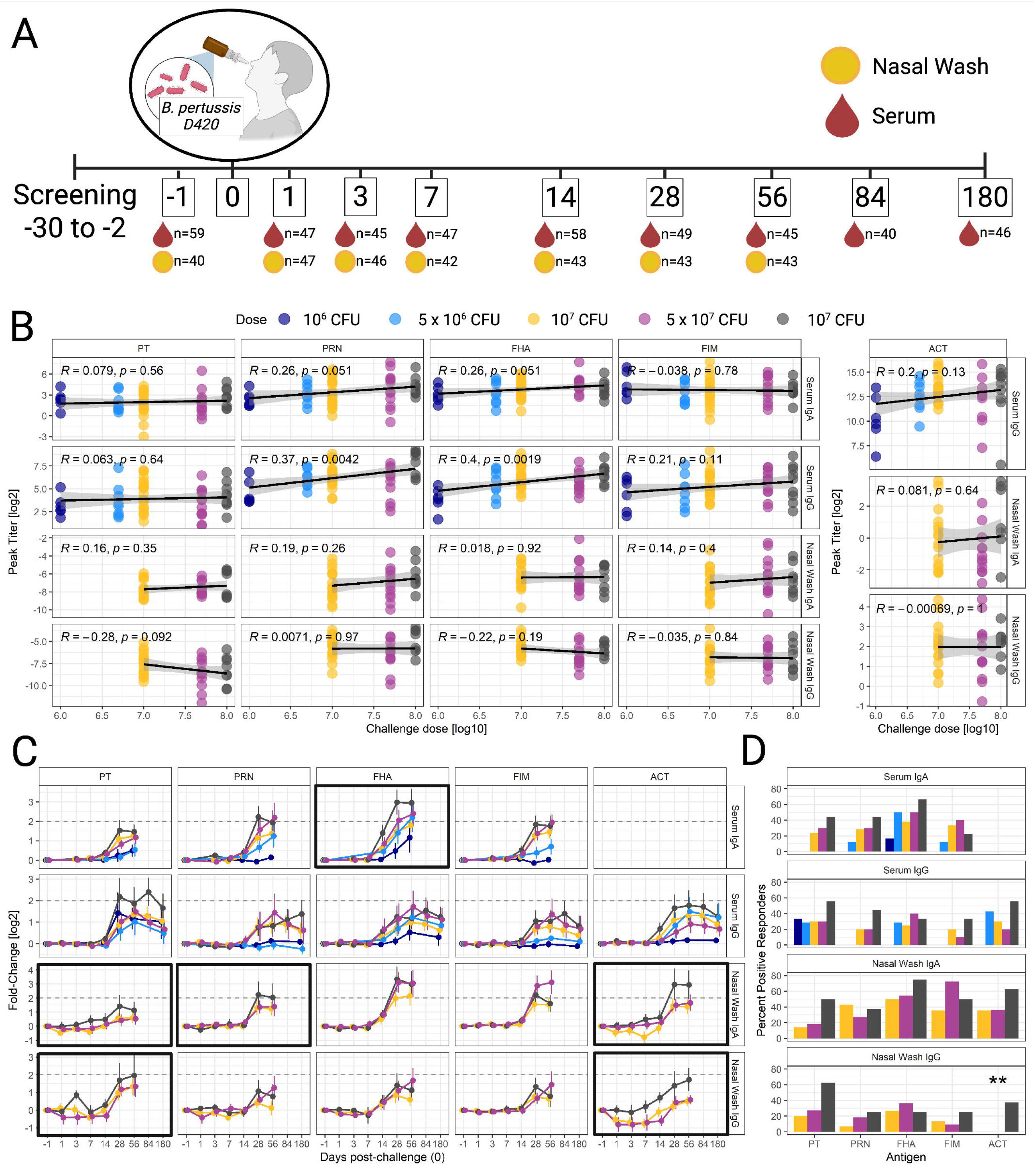
*B. pertussis* CHIM schematic representation and antibody responses post-challenge. **A**) Study participants were inoculated with *B. pertussis* D420 (Denoted as time 0). Inpatient observation spanned up to 21 days post-challenge, followed by 6 months of outpatient observation. Shedding was monitored during inpatient observation, and clinical disease outcome was monitored 6 days post-challenge. Blood and nasal tissue collected throughout the study were tested for the presence of pertussis antibodies, PT: Pertussis Toxin, PRN: Pertactin, FHA: Filamentous Hemagglutinin, FIM: Fimbriae 2/3, ACT: Adenylate cyclase toxin. **B**) Pearson correlation between challenge dose, log10(CFU), and peak antibody titer of each participant. **C**) Fold-change kinetics of antibodies, colored by inoculation dose. Bolded outlines represent a significant association with antibody kinetics based on mixed-effects modeling (**appendix pp. 7-10**). **D**) Positive response, defined as ≥4-fold from baseline. Asterisks represent Fisher’s test. ** p < 0.01.

The median age of the participants was 26 years (IQR: 23-33), and 43·4% (25/59) were women (**table 1**), and 78.0% (46/59) were white (**appendix pp. 6)**. Over a third of the participants (39·0% [23/59]) received aP as primary vaccine series, while the remaining 61·0% (36/59) received wP, and were evenly distributed across the dosage groups (Fisher test, p=0·31). When comparing wP- and aP-recipients, those who received an initial wP vaccine series were older than aP-recipients (median age 32 and 22, respectively; Wilcoxon, p<0·001), and more time had elapsed since their most recent pertussis vaccination (median 17·1 and 11·6 years, respectively; Wilcoxon, p<0·001) (**table 2**). The vaccine-priming stratification, reflecting childhood immunization, inherently captures age-related differences in exposure histories.

**Table 1.** Demographic characteristics of Cohorts, Data are n (%) or median (IQR)

|  |  |  |  |  |  |  |  |
| --- | --- | --- | --- | --- | --- | --- | --- |
|  | Total<br>n = 59 | 10<sup>6</sup> CFU<br>n = 6 | 5 x 10<sup>6</sup> CFU<br>n = 10 | 10<sup>7</sup> CFU<br>n = 22 | 5 x 10<sup>7</sup> CFU<br>n = 12 | 10<sup>8</sup> CFU<br>n = 9 | P-values |
| **Sex, Female**<br>**n (%)** | 25 (42.4%) | 2 (33.3%) | 4 (40.0%) | 8 (36.4%) | 7 (58.3%) | 4 (44.4%) | 0.80<sup>A</sup> |
| **Age, years**<br>**median**<br>**(IQR)** | 26.0<br>(23.0-33.0) | 32.5<br>(31.3-33.8) | 27.5<br>(26.0-31.3) | 24.0<br>(21.3-31.5) | 28.0<br>(23.8-34.0) | 25.0<br>(24.0-30.0) | 0.24<sup>B</sup> |
| **BMI**<br>**Median**<br>**(IQR)** | 25.4<br>(22.9-29.2) | 24.8<br>(22.0-33.3) | 25.1<br>(23.6-30.3) | 26.3<br>(23.7-30.5) | 24.7<br>(22.0-26.7) | 25.4<br>(23.5-27.4) | 0.79<sup>B</sup> |
| **Clinical Disease Outcome: n (%)** |  |  |  |  |  |  |  |
| **Non-Infected**<br>**Shedder** | 12 (20.3%)<br>47 (79.7%) | 3 (50.0%)<br>3 (50.0%) | 4 (40.0%)<br>6 (60.0%) | 3 (13.6%)<br>19 (86.4%) | 1 (8.3%)<br>11 (91.7%) | 1 (11.1%)<br>8 (88.9%) | 0.07<sup>A1</sup> |
| • Asymptomatic | • 14 (23.7%) | • 0 (0.0%) | • 4 (40.0%) | • 3 (13.6%) | • 4 (33.3%) | • 3 (33.3%) |  |
| • Symptomatic | • 33 (55.9%) | • 3 (50.0%) | • 2 (20.0%) | • 16 (72.7%) | • 7 (58.3%) | • 5 (55.6%) |  |
| **Original Pertussis Vaccine: n (%)** |  |  |  |  |  |  |  |
| **aP**<br>**wP** | 23 (39.0%)<br>36 (61.0%) | 1 (16.7%)<br>5 (83.3%) | 2 (20.0%)<br>8 (80.0%) | 11 (50.0%)<br>11 (50.0%) | 4 (33.3%)<br>8 (66.7%) | 5 (55.6%)<br>4 (44.4%) | 0.31<sup>A</sup> |
| **Years since previous reported pertussis vaccine** |  |  |  |  |  |  |  |
| **Median**<br>**(IQR)** | 14.1<br>(10.9-18.3) | 25.9<br>(17.3-32.9) | 14.1<br>(13.2-16.9) | 12.5<br>(8.4-18.7) | 16.7<br>(12.8-24.9) | 11.8<br>(11.1-15.0) | 0.05<sup>B</sup> |
| <sup>A</sup>: Fisher Test |  |  |  |  |  |  |  |
| <sup>A1</sup>: Fisher test between Non-infected, Asymptomatic, and Symptomatic |  |  |  |  |  |  |  |
| <sup>B</sup>: Kruskal-Wallis |  |  |  |  |  |  |  |

**Table 2.**
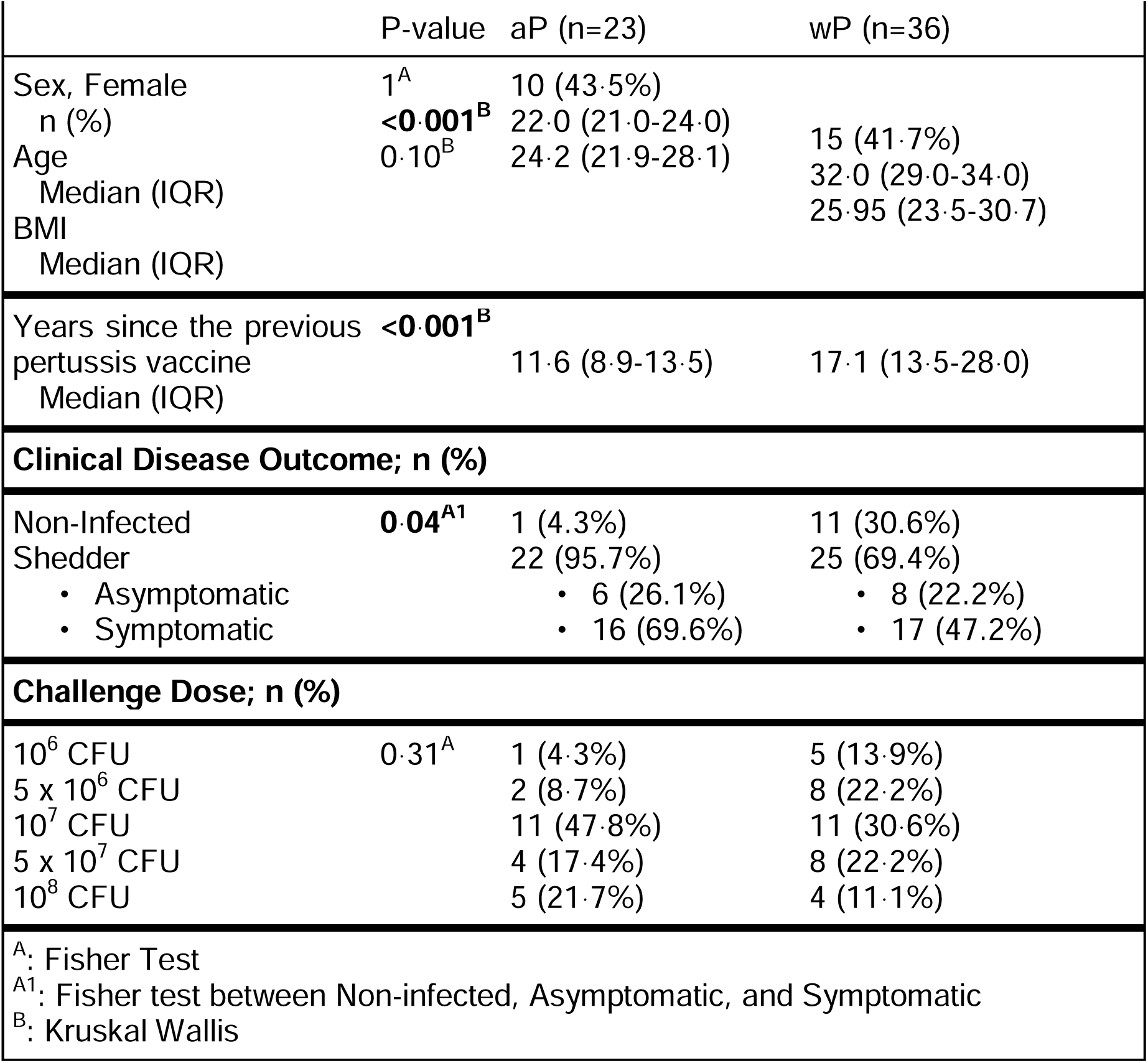
Acellular [aP] or whole-cell [wP] pertussis vaccines. Data are n (%) or median (IQR)

We first examined mucosal and serum antibodies elicited by *B. pertussis* in relation to the inoculation dose. High challenge doses were associated with higher peak titers of PRN (Pearson r=0·37; p=0·004) and FHA (r=0·40; p=0·002) serum IgG (**figure 1B).** There was also a weak positive correlation between inoculation dose and PRN and FHA serum IgA (r=0·26 for both; p=0·05 for both). Both serum and nasal antibodies, except for PT nasal IgA (p=0·11) and FIM serum IgA (p=0·05), exhibited significant non-linear temporal changes post-challenge (mixed-effects model, p<0·05) (**appendix pp 7-10**). The positive response rate (PRR) (defined as a ≥4-fold increase from baseline) was generally not significantly different between dose groups; only ACT nasal IgG showed a significant difference, with a 37·5% PRR in the 10^8^ CFU group (**figure 1D)**. Additionally, except for PT, nasal IgA outpaced nasal IgG in PRR across all challenge doses (**figure 1D)**. Most participants, regardless of dose group, exhibited positive nasal IgA responses to FHA. Similarly, the majority (>50%) of participants in the 5×10^6^, 5×10^7^, and 10^8^ dose groups had a positive FHA serum IgA response, while none of the groups reached >50% PRR for FHA IgG in either compartment (**figure 1D)**.

Bacterial shedding was defined as detection of *B. pertussis* starting at six days post-challenge, based on ≥1 positive culture or ≥3 positive PCRs. The majority (80%) of the participants evaluated for antibody responses shed *B. pertussis* post-inoculation, and the proportion of shedders increased with the challenge doses (Cochran–Armitage test for trend, Z=2·38, p=0·02), rising from 50% in the 10^6^ CFU cohort to approximately 90% in the 5×10^7^ and 10^8^ CFU cohorts (**table 1**). Clinical disease outcome was defined as: non-infected (no shedding), asymptomatic infection (shedding without respiratory symptoms), and symptomatic infection (shedding with respiratory symptoms). There was a significant difference in clinical disease outcome between wP and aP vaccine recipients (Fisher test, p=0·04), with the wP-recipient group having a higher rate of non-infected individuals (30·6% vs 4·3%) and a lower proportion of symptomatic infection (47·2% vs 69·6%) (**table 2**).

We next examined the relationship between pre-existing *B. pertussis* immunity and clinical outcome post-challenge, as well as the primary vaccine type. Partial least squares discriminant analysis (PLS-DA) was used to assess the discriminatory power of baseline humoral immunity in future disease outcomes (**figure 2A)**. Nine features contributed most strongly to this discrimination (Variable Importance in Projection [VIP] >1): FHA nasal IgA, FIM serum IgG, FIM nasal IgA, PRN nasal IgA, PT nasal IgA, PT nasal IgG, PT serum IgG, FHA serum IgA, and FIM nasal IgG. Non-infected participants had higher baseline FHA nasal and serum IgA than either asymptomatic or symptomatic participants, and higher FHA nasal IgG than symptomatic participants (Wilcoxon tests, p<0·05) **(figure 2B).** At baseline, non-infected participants also had higher PT nasal IgG titers than asymptomatic participants (p=0·03) and higher FIM serum IgA than symptomatic (p=0·009) or asymptomatic (p=0·01) participants. PLS-DA analysis was also used to compare baseline antibody levels between wP and aP recipients (**figure 2C)**. Eight features emerged as key discriminators of vaccine priming (VIP scores >1): PRN (nasal IgA and IgG, and serum IgA), PT (nasal IgA and IgG, and serum IgA and IgG), and FHA (nasal IgG). The wP recipients had higher baseline PRN nasal IgA and serum IgA (Wilcoxon, p=0·03 and 0·003, respectively), and FHA serum IgA (p=0·008) than aP recipients (**figure 2D).**

**Figure 2.**
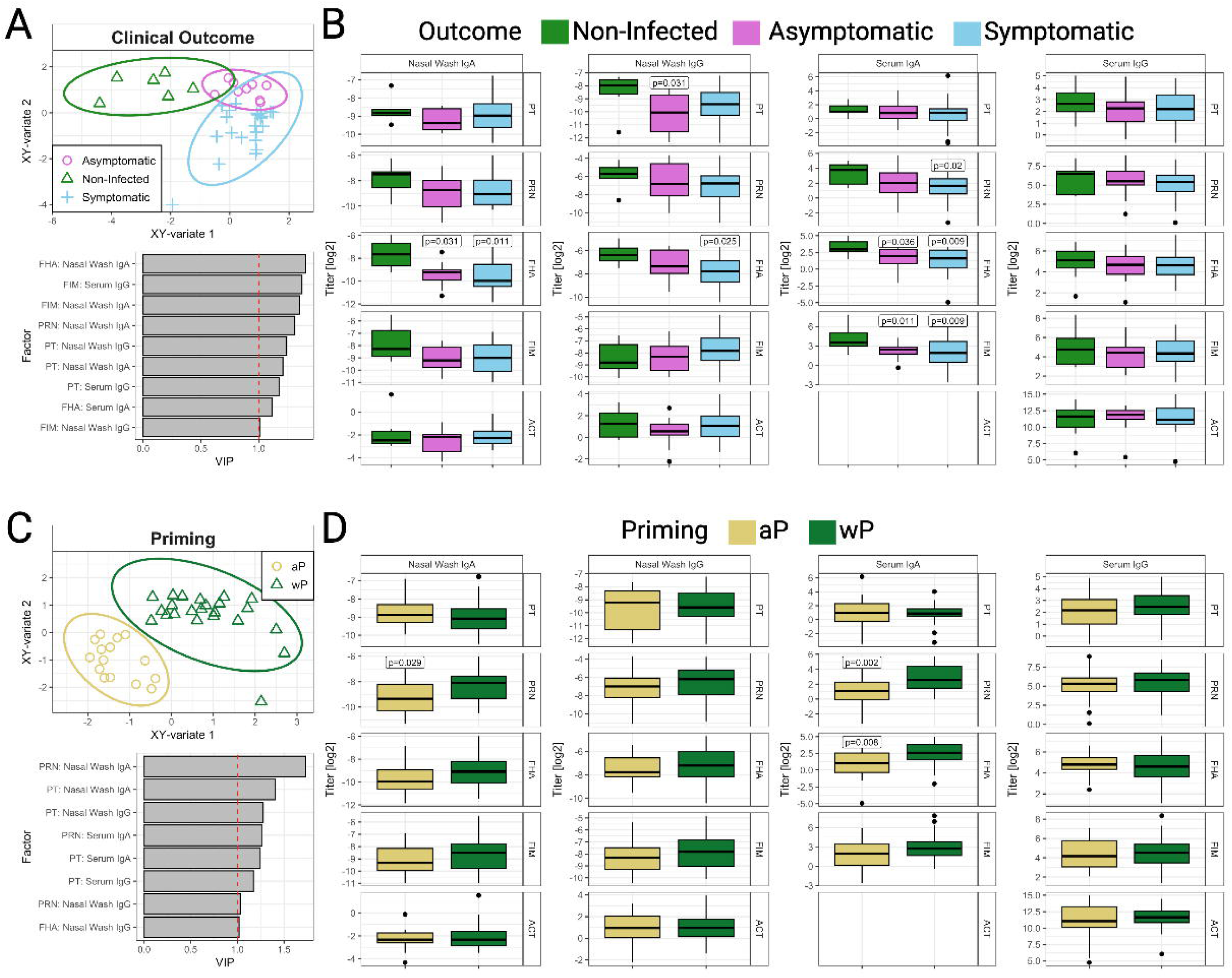
Pertussis mucosal and systemic antibodies at baseline. **A**) Top: Partial Least Squares Discriminant Analysis (PLS-DA) of baseline titers and clinical outcome. Bottom: Variable Importance in Protection (VIP) features. **B**) Baseline titers with Wilcoxon p-values comparing individuals with different clinical disease outcomes vs the non-infected (reference group). **C**) Top: PLSDA of baseline titers comparing individuals who received acellular pertussis (aP) or whole cellular pertussis vaccine (wP) as primary vaccine series. Bottom: VIP features. **D**) Baseline titers comparing aP- and wP-primed individuals. P-values represent significant Wilcoxon p-values. PT: Pertussis Toxin, PRN: Pertactin, FHA: Filamentous Hemagglutinin, FIM: Fimbriae 2/3, ACT: Adenylate cyclase toxin.

The temporal evolution of pertussis bacterial load was monitored via nasal wash CFUs and evaluated through day 16 post-challenge, at which time azithromycin was generally given (average day administered: 16·02±1·91) (**figure 3A-C)**. Non-infected participants had no bacterial shedding at any time point, indicating the absence of residual colonies from the challenge dose (**figure 3A).** Those who shed *B. pertussis* had a peak bacterial load at 9·0±3·8 days post-challenge; there was no significant difference in timing of peak bacterial load between asymptomatic (Peak day: 10·14±3·25) and symptomatic (8·55±3·93) participants (Wilcoxon p=0·15) **(figure 3A)**. There was a trend toward higher bacterial load across time points for the different dose groups, with recipients of the 10^8^ CFU (highest) challenge dose having the highest bacterial load (**figure 3B).** Starting six days post-challenge, aP recipients had a significantly higher bacterial load than wP recipients (Wilcoxon, p<0·05), and this difference expanded thereafter (**figure 3C**).

**Figure 3.**
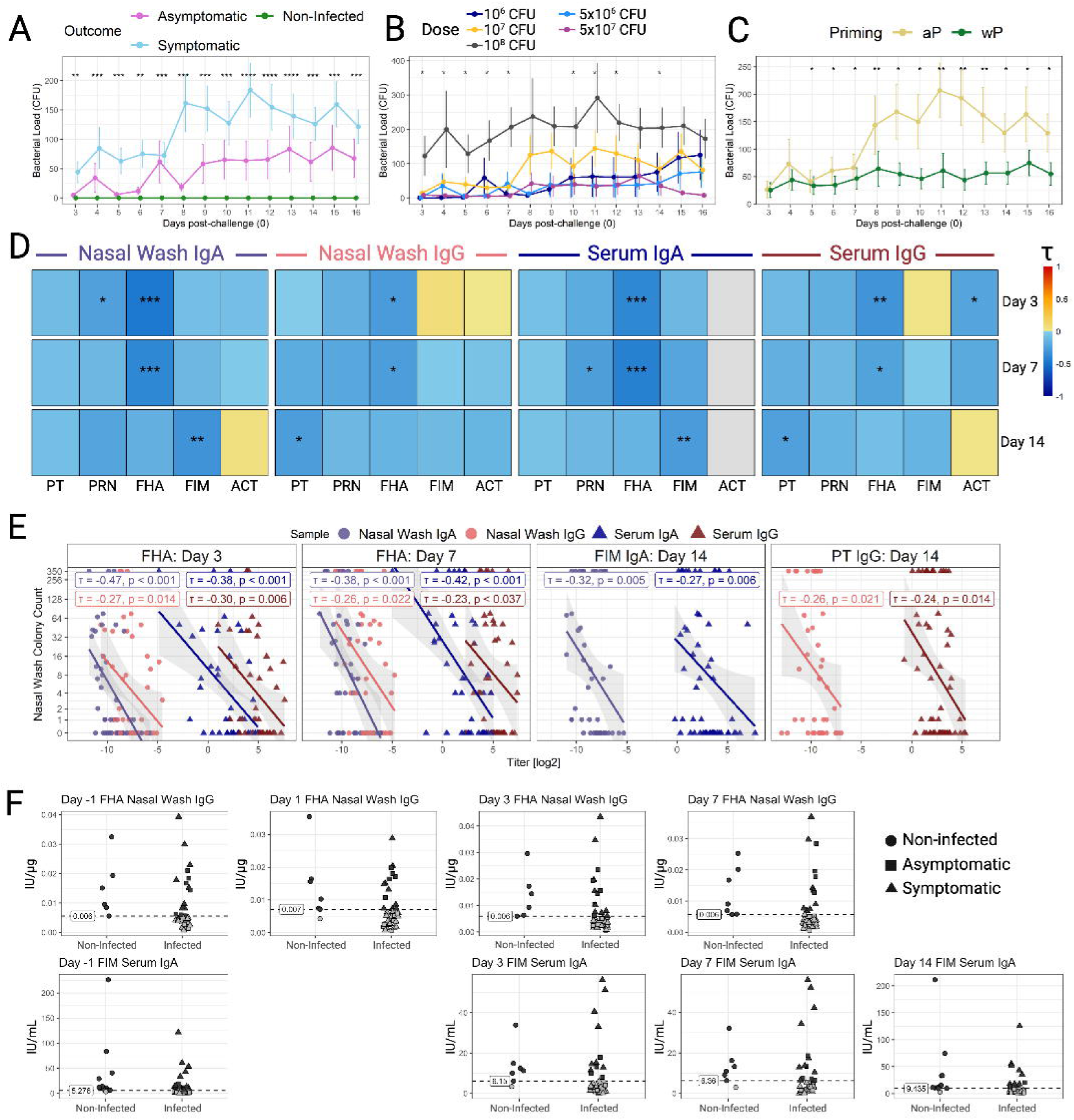
Correlation between antibody titers and nasal bacterial shedding post-challenge. Pertussis colony-forming unit (CFU) from nasal wash post-challenge comparing (**A**) clinical disease outcome, (**B**) different inoculation doses, and (**C**) recipients of aP and wP primary vaccine series. On day 16, azithromycin was administered to all patients unless previously administered when symptoms developed. **D**) Kendall’s tau correlation between antibody titers (nasal IgA, nasal IgG, serum IgA, and serum IgG) and bacterial shedding post-challenge. Colors represent the correlation coefficient, and the asterisks indicate the significance of the corresponding correlation. *** p < 0·001, ** p < 0·01, * p < 0·05. **E**) Plots featuring significant associations between antibody titers and bacterial shedding in multiple sample types. **F**) Discrimination thresholds (infected vs. non-infected) identified through AUC and ROC analysis. Thresholds for discriminatory performance were defined as AUC≥0·70, Youden index≥0·30, sensitivity>0·70, and specificity>0·50 (**appendix 10-13**). FHA and FIM antibodies are shown, as they were identified as disease-discriminatory features at multiple time points. PT: Pertussis Toxin, PRN: Pertactin, FHA: Filamentous Hemagglutinin, FIM: Fimbriae 2/3, ACT: Adenylate cyclase toxin.

The association between pertussis bacterial load and antibody levels was investigated at days 3, 7, and 14, time points for which both clinical and immunological data were available. Kendall’s tau-b correlation was used, which is appropriate for non-normally distributed data and accounts for the large proportion of zero counts in nasal shedding (**figure 3D)**. At days 3 and 7 post-challenge, FHA IgA and IgG in both serum and nasal wash were negatively correlated with bacterial load (Kendall’s τ=-0·23 to -0·47, p<0·05) (**figure 3E**). At day 14, negative correlations were also observed for PT IgG in nasal wash (τ=-0·26, p=0·02) and serum (τ=-0·24, p=0·014), as well as FIM IgA in nasal wash (τ=-0·32, p=0·005) and serum (τ=-0·27, p=0·006) (**figure 3E**). Receiver operating characteristic (ROC) analysis with leave-one-out cross-validation was used to evaluate the ability of individual antibodies to discriminate between shedders and non-shedders, and to evaluate the corresponding antibody thresholds **(appendix pp. 11-14)**. Two antigens, FHA nasal IgG and FIM serum IgG, were identified as discriminators at multiple time points through 2 weeks post-challenge (**figure 3F)**; no significant discriminators were observed across any antigens or sample types on days 28 and 56 post-challenge. Nasal FHA IgG demonstrated infected/non-infected discriminatory capacity from baseline through day 7 post-challenge, with an optimal threshold of 0·006 IU/µg that remained relatively stable over time **(figure 3F).** FIM serum IgA also exhibited discriminatory capacity, with the threshold increasing from 5·28 at baseline to 9·44 IU/µg on day 14.

There was a strong correlation between serum and nasal IgG levels across FHA, PRN, and FIM (Pearson r: 0·81 to 0·95) and a modest correlation between serum and nasal IgA (Pearson r: 0·44 to 0·72) (**appendix pp. 15)**. The strength of IgG correlations for PT improved over time, with Pearson’s r values increasing from 0·67 on day 1 to 0·81 on day 56. Similarly, serum and nasal PT IgA correlations only became significant 28 days post-challenge.

Unlike infected participants, those who remained uninfected exhibited little to no change in serum and mucosal antibodies post-challenge and did not have a positive response for any antigen (**figure 4A)**. Asymptomatic participants exhibited earlier and more robust nasal responses to PT than symptomatic participants, whereas symptomatic participants exhibited more robust PT serum responses several weeks post-challenge (Wilcoxon, p<0·05). Symptomatic participants also had later and more robust serum IgG responses to PRN and FIM than asymptomatic participants (p<0·05). Symptomatic and asymptomatic participants exhibited higher PRR for FHA serum IgA (0·0%, 63·3%, 35·7% for non-infected, symptomatic, and asymptomatic participants, respectively; Fisher’s p<0·001), FHA serum IgG (0·0%, 36·7%, 28·6%; p=0·04), FHA nasal IgA (0·0%, 68·2%, 40·0%; p=0·02). Additionally, PT serum IgG (0·0%, 43·3%, and 35·7%; p=0·02), ACT serum IgG (0·0%, 43·3%, and 21·4%; p=0·015), and FIM serum IgA (0·0%, 43·3%, and 35·7%; p=0·04), also showed higher PRR in infected participants. Nasal and serum FHA IgA were among the top four features discriminatory of clinical outcomes on days 3 and 7 post-challenge, but their importance diminished by day 14 (VIP<1) (**figure 4B**). In contrast, nasal and serum PT IgG emerged as the primary discriminators on days 14 and 56 post-challenge, with the VIP increasing from 1·06 on day 3 to 1·58 on day 56. This outcome-discriminatory pattern suggests a time-dependent antibody-associated resolution of illness, with an early critical involvement of mucosal FHA antibodies followed by a later contribution from systemic PT antibodies.

**Figure 4.**
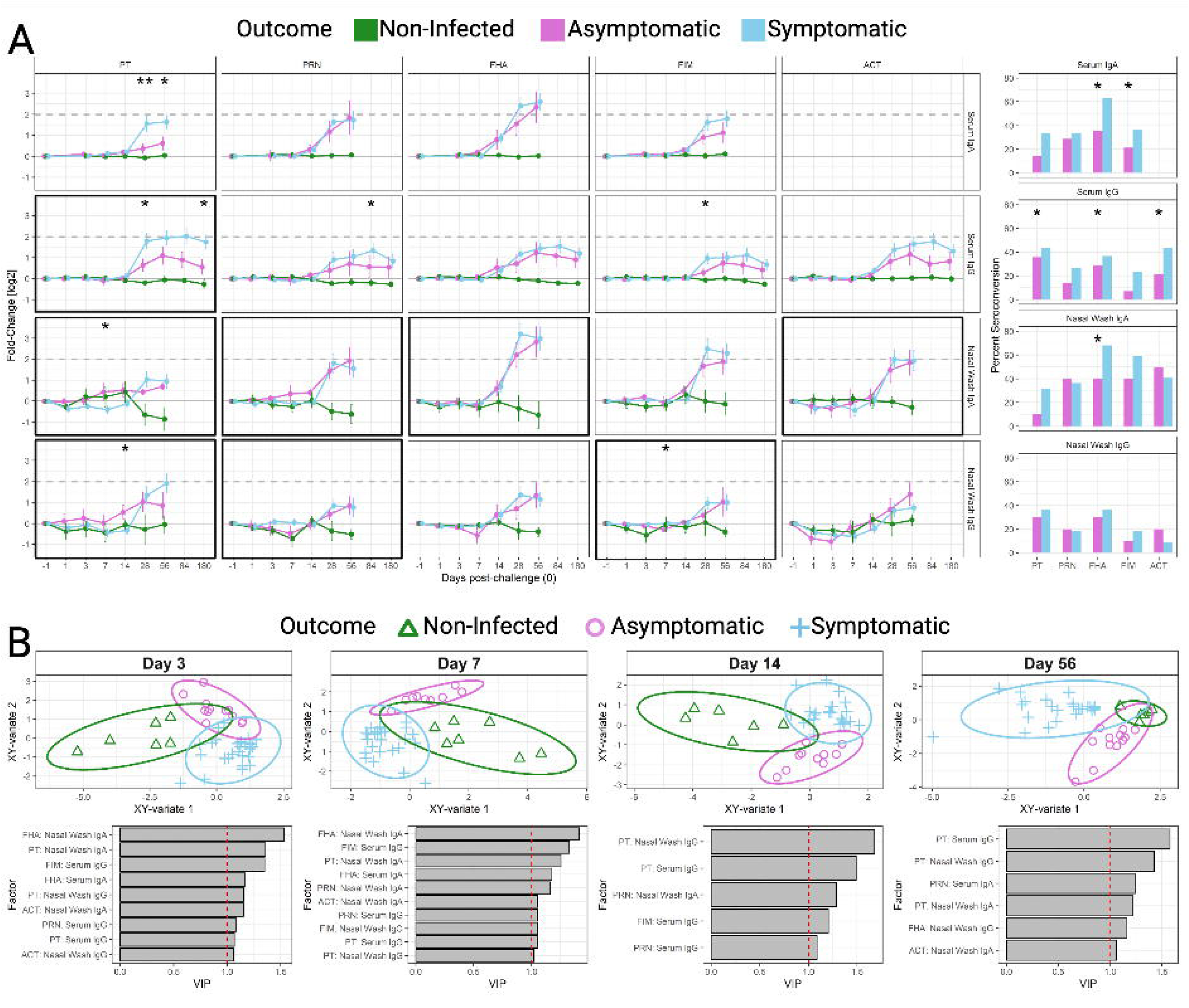
Systemic and mucosal antibody titers by clinical disease outcome. **A**) (Left) Fold-change kinetics of antibodies colored by clinical disease outcome. Non-infected: No reported shedding (PCR or colony count). Symptomatic: Shedding with respiratory symptoms. Asymptomatic: Shedding without respiratory symptoms. Bolded outlines represent significant associations with antibody kinetics based on mixed-effects modeling (**appendix pp. 7-10**). (Right) Percent positive response (≥4-fold change) at any time point post-challenge. Asterisks indicate the Fisher test for positive response. * p < 0·05. **B**) (Top) Partial least squares discriminant analysis (PLS-DA) between clinical disease outcome groups at day 3, 7, 14, and 56. (Bottom) Variable Importance in Projection (VIP) features.

aP recipients exhibited stronger responses at later time points (≥28 days post-challenge), including PRN and FHA serum IgA and IgG, FIM serum IgA, PT serum IgG, and ACT serum IgG (Wilcoxon p<0·05) (**figure 5A)**. aP recipients had higher PRR than wP recipients for PT serum IgG (55·5% and 20·0%, respectively; Fisher’s p=0·015) and ACT serum IgG (50·0% and 17·1%; p=0·01) (**figure 5A).** PT IgA (nasal and serum) was a discriminating factor on days 3, 7, and 14 (**figure 5B).** On day 56, most VIP factors were serum IgG, except for FHA and PRN serum IgAs, which were VIPs at all time points.

**Figure 5.**
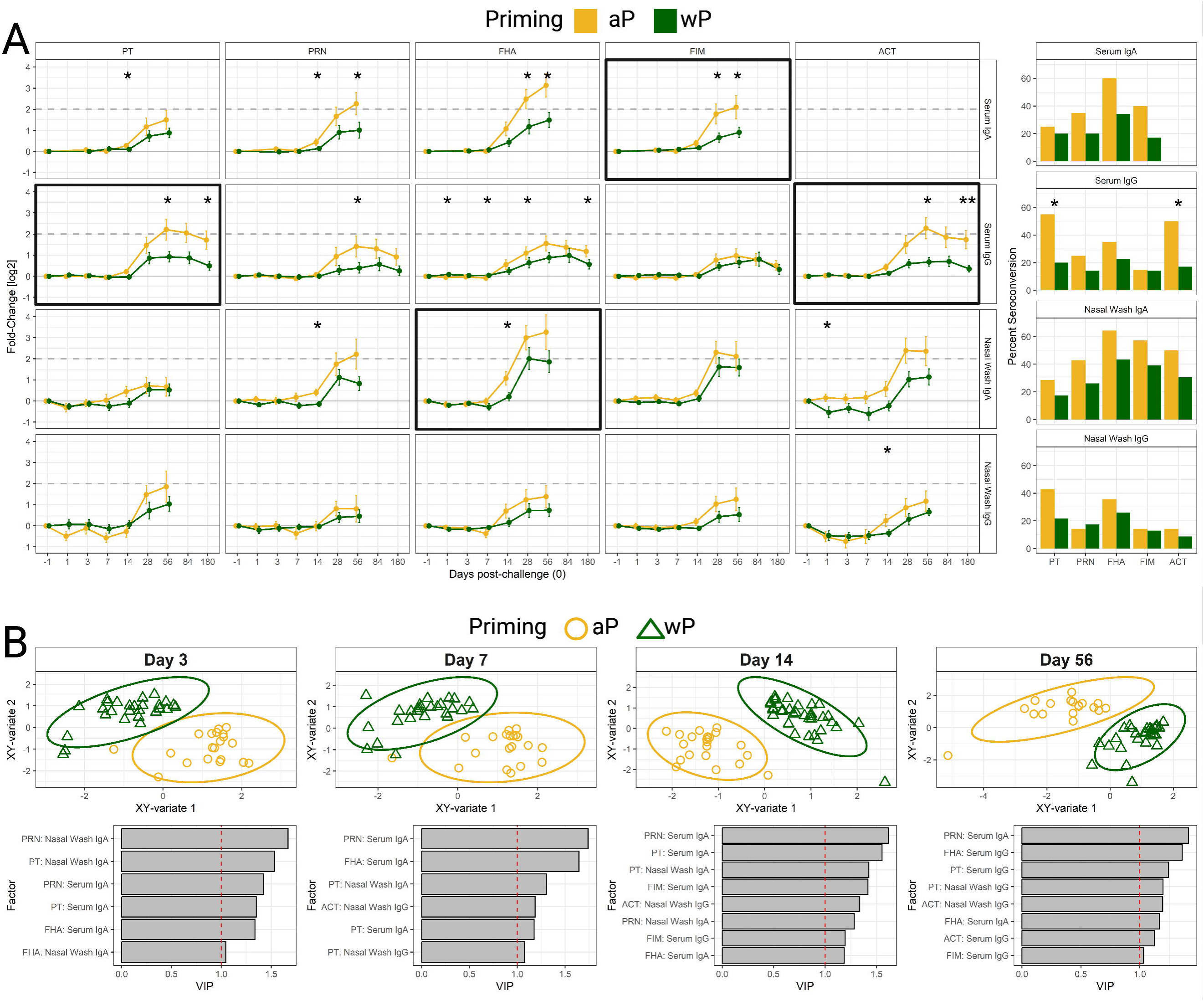
Systemic and mucosal antibody titers by primary vaccine type. **A)** (Left) Fold-change kinetics of antibodies, colored by priming vaccine type: acellular pertussis vaccine (aP) or whole-cell pertussis vaccine (wP). Bolded outlines represent significant associations with antibody kinetics based on mixed-effects modeling (**appendix pp 7-10).** (Right) Percent positive response (≥4-fold change) at any time point post-challenge. Asterisks indicate the Fisher test for positive response. * p < 0·05. **B**) (Top) Partial least squares discriminant analysis (PLS-DA) between vaccine priming groups at day 3, 7, 14, and 56. (Bottom) Variable Importance in Projection (VIP) features.

## DISCUSSION

This study uniquely describes a longitudinal quantitative assessment of mucosal and systemic antibodies specific to major pertussis antigens, and a correlational analysis to bacterial shedding and clinical disease outcomes in individuals who participated in a pertussis dose-escalation CHIM study. Important insights emerged, hinting at a protective role of mucosal FHA IgG and IgA and FIM2/3 serum IgA early after infection, with a subsequent contribution of serum and mucosal PT IgG, and at a long-lasting imprinting of host immunity by primary vaccination, with aP conferring weaker immunity and having a higher rate of symptomatic infection.

While the proportion of individuals shedding organisms increased with the inoculation dose, their antibody response rate (≥4-fold above baseline) remained unaffected. A majority of participants had positive serum and nasal IgA FHA responses, whereas a smaller number exhibited an IgG response. Through multiple analytical approaches, FHA nasal antibodies emerged as a strong discriminator between infected and non-infected individuals. At the outset of the study, FHA IgA was significantly higher in participants who remained non-infected than in those who would become infected, regardless of symptom status. During the first week post-challenge, mucosal and systemic FHA antibodies remained negatively associated with *B. pertussis* bacterial load, and nasal FHA IgG titers below 0.006 IU/µg were associated with infection.

FHA is an adhesion protein that facilitates *B. pertussis* attachment to epithelial cells. It is also an immunomodulatory protein that facilitates uptake by macrophages and promotes an anti-inflammatory response by inducing IL-10 and IL-6, and suppressing IL-12, helping the bacteria evade early immune clearance.^11,12^ Pertussis FHA also promotes bacterial aggregation and biofilm development; hence, the need to block its effects. FHA-IgA is a predominant host mucosal effector against pertussis.^13^ FHA antibodies play a critical role in protective immunity; they have been found to prevent infection in mice and to be elevated in children who remained healthy during a pertussis outbreak.^14^ Our correlative analysis supports a synergistic role for mucosal and systemic FHA antibodies in preventing *B. pertussis* colonization of the respiratory airways soon after infection.

FIM 2/3 serum and nasal IgA were negatively correlated with bacterial load on day 14. The seroconversion rate for serum FIM IgA differed significantly across clinical disease outcomes (non-infected, symptomatic, and asymptomatic). Interestingly, it did not appear as a VIP for clinical disease outcomes at any time point, although antibody titers reliably discriminated disease status. FIM serum IgA and IgG have been shown to reduce *B. pertussis* attachment to a human alveolar epithelial cell line via agglutination.^15^ The tracking of FIM IgA with reduced bacterial shedding suggests a similar antibody-mediated mechanism of bacterial clearance during human infection. While mucosal IgA could directly block cell attachment and prevent infection, the role of serum IgA is less intuitive. Whether systemic IgA acts independently or reflects parallel mucosal production and function remains to be investigated.

PT serum and nasal IgG were also negatively correlated with bacterial load on day 14, and these were the two most important discriminators of pertussis clinical outcome from day 14 onwards. Because participants were excluded from the study if they had >20 EU/mL of PT IgG in serum, the study could not determine whether high PT antibody levels could have prevented infection. Regardless, it is noteworthy that late PT serum IgG responses (day 28 and 180) were stronger in symptomatic than in asymptomatic individuals, suggesting that PT humoral immunity is associated with more severe infection.

The majority of participants who remained uninfected were childhood wP recipients. Their protective immune status is noteworthy despite being older and having a longer interval since vaccination than the aP participants. aP- and wP-primed individuals had distinct baseline immune profiles that persisted after challenge. This observation highlights the long-lasting effects of early vaccine-induced immune imprinting and the importance of effective immunization in establishing a robust, protective immune trajectory. For example, aP recipients exhibited more robust systemic PT and ACT IgG responses starting 56 days post-challenge, consistent with their higher infection rate. In animal models, aP vaccination inhibited severe pertussis symptoms but did not reduce colonization or shedding.^16^

There is a paucity of data on pertussis immunity in human mucosal tissues. FHA IgA was detected in saliva after the onset of pertussis symptoms and in a majority of patients during late convalescence, whereas PT IgA was absent.^17^ Similarly, nasal FHA IgA increases (1·5-to 2-fold) were reported in recipients of an intranasal wP vaccine, while PT IgA was not detected.^18^ Intriguingly, in our study, baseline nasal PT IgG titers were higher in non-infected as compared to infected, asymptomatic individuals. These observations highlight the contrasting responses evoked by different antigens, involving antibodies with distinct functional capabilities in mucosal and systemic compartments.

In a recent CHIM, the *B. pertussis* strain B1917 (NCT03751514) was administered at doses ranging from 10^3^ to 10^5^ CFU, only to individuals who received the wP vaccine in childhood.^6,7^ Similar to our findings, colonization rates increased with the challenge dose, reaching 80% in the 10^5^ CFU cohort. This study examined only serum IgG titers and reported significant negative correlations between FHA, PRN, and FIM antibodies and maximum CFU counts.^7^ However, no differences in baseline titers were found between colonized and uncolonized individuals.^6^ Consistent with our observed discrimination of clinical outcome by baseline titers, a subsequent CHIM focused on BPZE1 vaccine efficacy also reported higher initial serum FHA antibody levels in uncolonized vaccine recipients.^19^ While association analyses do not prove causality, overwhelming evidence from this and other studies supports FHA and PT as putative protective antigens that should be included in vaccine candidates.

The antimicrobial function of antibodies is not considered in our analysis. Measurement of antibodies mediating pertussis toxin neutralization and opsonophagocytic killing in serum and mucosal secretions from challenged volunteers is underway. Importantly, excluding individuals with PT serum IgG >20 EU/mL precluded analysis of the role of high systemic PT antibody levels in preventing disease. Lastly, the relatively small sample size may affect the stability of performance estimates and limit the generalizability of these results. Validation of these results in a larger and more diverse population would be desirable.

In summary, we demonstrate a correlational association between FHA, FIM 2/3, and PT mucosal and systemic antibodies and reduced *B. pertussis* colonization, as well as the impact of prior vaccine history on pertussis clinical outcomes and post-challenge responses. Our results also revealed a coordinated, temporally associated mucosal and systemic protective immunity against *B. pertussis* infection.

## Data Availability

The deidentified compiled dataset and analysis scripts used for this publication are available at github.com/PMilletich/Pertussis_CHIM_BindingAntibodies

## Contributors

PLM: Formal Analysis, Visualization, Writing - Original Draft, Writing - Reviewing & Editing. ME: Writing - Reviewing & Editing. KM: Investigation, Methodology, Writing - Original Draft. AC: Investigation and Project Administration. SH^1^: Writing - Reviewing & Editing. SH^2^: Writing - Reviewing & Editing. MFP: Funding Acquisition, Supervision, Writing - Original Draft, Writing - Reviewing & Editing

