## Appendix for "Mucosal and Systemic Antibodies Associated with Clinical Protection in a Pertussis Controlled Human Infection Model"

Supplementary Materials

### 1 Methods

#### 1.1 Study Design

This controlled human infection trial was a dose-escalation study in which participants received a single intranasal B. pertussis 420 inoculation at seven doses: 10^4^, 10^5^, 5x10^5^, 106, 5x10^6^, 10^7^, 5x10^7^, 10^8^ colony-forming units (CFU). All participants received a 5-day course of azithromycin eradication therapy either 24-48 hours after developing symptoms or at the end of the 16-day post-challenge observation period.

#### 1.2 Participants

Healthy males and non-pregnant females between 18 and 40 years of age, inclusive, were recruited at the Canadian Center for Vaccinology, Halifax, Nova Scotia, Canada. Participants were excluded if they were vaccinated with pertussis within the last 5 years, had more than seven cumulative pertussis vaccine Cohorts from infancy to date of screening, had a history of laboratory-confirmed pertussis infection, had an antibody titer to pertussis toxin > 20 EU/mL at screening, or had nasopharyngeal detection of B. pertussis prior to challenge using culture isolation and/or PCR detection. The complete eligibility criteria are published at https://clinicaltrials.gov/study/NCT05136599.

Participants were non-randomized and assigned sequentially to escalating inoculation Cohorts. No masking was done (Open Label).

#### 1.3 Clinical Disease Outcomes

Participants were classified as positive for pertussis if they had one or more positive cultures or two or more positive PCRs six days post-challenge. Symptomatic participants were positive for pertussis and had 2 or more symptoms, including at least 1 respiratory symptom (cough or sneeze). Asymptomatic participants were positive for pertussis but had no respiratory symptoms. Non-infected participants tested negative for pertussis.

#### 1.4 Laboratory Assays

##### 1.4.1 Bordetella pertussis Multiplexed Binding Antibody Assay

Briefly, pre-coated (Filamentous Hemagglutinin, Fimbriae 2/3, Pertussis Toxin, Pertactin, Tetanus Toxoid, and Diphtheria Toxoid) microtiter plates were removed from 4°C storage and allowed to equilibrate to room temperature (RT), then blocked with 150 μL/well of blocking buffer (1X PBS-T with 10% nonfat dry milk [NFDM]) and incubated at 22°C in an incubator for 1 h while shaking at 400 rpm. Samples, standards, and controls were diluted in PBS-T + 10% NFDM. Plates were washed using a BioTek™ 405LS plate washer, which performs three washes, rotates the plate 180°, and washes three additional times (six total). 50 μL per well of each diluted sample, negative control, and assay-specific positive control (serum or nasal wash) were added in duplicate, along with a 2-fold serial dilution of the standard. After adding samples, controls, and standards, the plates were incubated at 22°C in an incubator for 1 h with shaking at 400 rpm. The secondary antibody (either IgG or IgA) was diluted in assay diluent, added at 50 μL per well following a wash, and incubated under the same conditions. After a final wash step, 150 μL of MSD Gold Read Buffer A was added to all wells, and plates were immediately read using the MSD QuickPlex SQ 120. Data were analyzed using MSD Discovery Workbench software. Standard curves were generated by fitting the electrochemiluminescence (ECL) signals from the standards to a four-parameter logistic (4PL) model with a 1/Y² weighting. Ab concentrations in controls and samples were determined by backfitting their ECL signals to the RS1 standard curve and multiplying by the sample’s dilution factor.

##### 1.4.2 ACT assay

A 96-well flat-bottom Immulon 2HB (Thermo Labsystems #3455) microtiter plates were coated with 0.5 μg/mL of recombinant adenylate cyclase toxin (ACT) diluted in 1X phosphate-buffered saline (PBS), pH 7.4, and incubated overnight at 4°C. Plates were washed using the BioTek™ 405LS plate washer, three washes with 1X PBS + 0.05% Tween-20 [PBST], followed by a 180° rotation and three additional washes. Plates were then blocked with 220 μL/well of blocking buffer (PBST + 5% nonfat dry milk [NFDM]) and incubated at 37°C for 1 h. After washing again as described above, 100 μL/well of diluted In-House Standard, assay-specific controls (serum or nasal wash), and test samples (prepared in PBST + 5% NFDM) were added to the plates in duplicate and incubated at 37°C for 1 h. Plates were washed again as described, and 100 μL/well of horseradish peroxidase (HRP)-conjugated secondary antibody (IgG or IgA), diluted in blocking buffer, was added and incubated for 1 h at 37°C. Plates were washed again as described, then 100 μL of tetramethylbenzidine (TMB) substrate was added to each well. Color development was carried out for 15 min in the dark on a plate rotator. The reaction was stopped by adding 100 μL/well of 1M phosphoric acid, and absorbance was immediately measured at 450 nm (OD_450_). Raw data (OD_450_) were analyzed by 4PL regression, and anti-ACT Ig concentrations were interpolated from the In-House Standard curve in GraphPad Prism.

##### 1.4.3 Total Mucosal Antibody Concentration Assay

Isotyping Panel 1 Human/NHP Calibrator blend (#C0203-2) was diluted (1:20 then serially diluted 4-fold), and nasal wash samples were diluted 1:5000, with kit diluent (#R50AA-3). 150 µL/well of Blocker A solution (#R93BA-2) was added to an Isotyping Panel 1 (Human/NHP) Plate (#N45203A-1) and incubated at RT with shaking (700 rpm) for 30 min. Plates were washed three times with wash buffer (1X PBS-Tween (0.05%)). After washing, 25 µL of samples, controls (neat), and calibrator were added to the plate, which was then incubated at RT with shaking for 2 h.  After incubation, the plates were washed three times with wash buffer, and then 25 µL of 1X detection antibody working solution (SULFO-TAG anti-Human/NHP IgA (#D20JJ-3), SULFO-TAG anti-Human/NHP IgG Antibody (#D20JL-3), and diluent) was added to each well. The plates were incubated at 22°C, shaking, for 2 h, then washed three times with wash buffer. 150µL MSD 2X Read Buffer T (#R92TC-3) was added to each well, and plates were immediately read using MSD QuickPlex SQ 120. A calibration curve was established by fitting the calibrator signals to a 4PL model with a 1/Y2 weighting. Antibody concentrations (AU/mL) were then determined by backfitting the ECL signals to the calibration curve.

##### 1.4.4 Mucosal antibody normalization

Antibody titers for all antigens (EU/mL for ACT and IU/mL for all others) in nasal wash samples were normalized by dividing the specific antibody response by the total antibody concentration (µg/mL) for each sample. The resulting normalized titers were used in all analyses for this study.

#### 1.5 R packages

mixomics package (version 6.32.0), RVAideMemoire package (version 0.9.83.12).  lmerTest (version 3.1-3) and splines (version 4.3.3). pheatmap (version 1.0.12).

#### 1.6 Linear mixed-effect modeling

Fold-change trajectories over time for each sample type and pertussis-specific antibody were analyzed through a linear mixed-effect model. Time was modeled using a natural cubic spline with three degrees of freedom to allow modeling of potentially non-linear changes over time. Fixed-effects included scaled inoculation CFU dose, priming group, and disease outcome, each interacted with the spline-transformed time variable to evaluate differences in temporal response patterns. Participant ID was used as a random intercept to account for repeated measurements within individuals.

### 2 Supplementary Tables

#### Table S1. Ethnicity/Race between Dose Cohorts

| Ethnicity | Total | 10^6^ CFU | 5 x 10^6^ CFU | 10^7^ CFU | 5 x 10^7^ CFU | 10^8^ CFU |
| --- | --- | --- | --- | --- | --- | --- |
| White | 46 (78·0%) | 6 (100·0%) | 7 (70·0%) | 17 (77.3%) | 9 (75·0%) | 7 (77·8%) |
| Multiple | 5 (8·5%) |  | 2 (20·0%) | 1 (4·5%) | 1 (8·3%) | 1 (11·1%) |
| Black | 2 (3·4%) |  |  |  | 1 (8·3%) | 1 (11·1%) |
| Filipino | 2 (3·4%) |  | 1 (10·0%) | 1 (4·5%) |  |  |
| Arab | 1 (1·7%) |  |  | 1 (4·5%) |  |  |
| Chinese | 1 (1·7%) |  |  |  | 1 (8·3%) |  |
| Latin American | 1 (1·7%) |  |  | 1 (4·5%) |  |  |
| Western European | 1 (1·7%) |  |  | 1 (4·5%) |  |  |

#### Table S2. Mixed-effect model of Serum IgA

| Antigen | Factor | P-value | Sum Sq | Mean Sq | DF | DenDF | F-value |
| --- | --- | --- | --- | --- | --- | --- | --- |
| PT | CFU_Scale | 0·946 | 0·150 | 0·15 | 1 | 235·273 | 0·005 |
|  | ns(Day, df = 3) | 0·015 | 349·612 | 116·537 | 3 | 239·119 | 3·546 |
|  | Priming | 0·957 | 0·095 | 0·095 | 1 | 235·336 | 0·003 |
|  | Outcome | 0·981 | 1·232 | 0·616 | 2 | 236·009 | 0·019 |
|  | CFU_Scale:ns(Day, df = 3) | 0·939 | 13·346 | 4·449 | 3 | 236·232 | 0·135 |
|  | ns(Day, df = 3):Priming | 0·235 | 140·874 | 46·958 | 3 | 236·67 | 1·429 |
|  | ns(Day, df = 3):Outcome | 0·08 | 375·999 | 62·666 | 6 | 237·804 | 1·907 |
| PRN | CFU_Scale | 0·932 | 1·712 | 1·712 | 1 | 242·829 | 0·007 |
|  | ns(Day, df = 3) | 0·001 | 3728·892 | 1242·964 | 3 | 243·608 | 5·347 |
|  | Priming | 0·997 | 0·003 | 0·003 | 1 | 242·899 | <0·001 |
|  | Outcome | 0·998 | 0·826 | 0·413 | 2 | 243·575 | 0·002 |
|  | CFU_Scale:ns(Day, df = 3) | 0·655 | 377·053 | 125·684 | 3 | 240·698 | 0·541 |
|  | ns(Day, df = 3):Priming | 0·998 | 8·606 | 2·869 | 3 | 240·999 | 0·012 |
|  | ns(Day, df = 3):Outcome | 0·196 | 2021·637 | 336·939 | 6 | 242·359 | 1·449 |
| FHA | CFU_Scale | 0·898 | 2·176 | 2·176 | 1 | 190·803 | 0·017 |
|  | ns(Day, df = 3) | <0·001 | 4559·531 | 1519·844 | 3 | 238·133 | 11·548 |
|  | Priming | 0·999 | <0·001 | <0·001 | 1 | 190·902 | <0·001 |
|  | Outcome | 0·991 | 2·494 | 1·247 | 2 | 191·783 | 0·009 |
|  | CFU_Scale:ns(Day, df = 3) | <0·001 | 2683·056 | 894·352 | 3 | 235·668 | 6·796 |
|  | ns(Day, df = 3):Priming | 0·197 | 621·127 | 207·042 | 3 | 235·926 | 1·573 |
|  | ns(Day, df = 3):Outcome | 0·287 | 977·736 | 162·956 | 6 | 237·144 | 1·238 |
| FIM | CFU_Scale | 0·996 | 0·011 | 0·011 | 1 | 238·368 | <0·001 |
|  | ns(Day, df = 3) | 0·053 | 3164·167 | 1054·722 | 3 | 243·314 | 2·593 |
|  | Priming | 0·921 | 3·980 | 3·980 | 1 | 238·442 | 0·010 |
|  | Outcome | 0·997 | 2·649 | 1·324 | 2 | 239·154 | 0·003 |
|  | CFU_Scale:ns(Day, df = 3) | 0·904 | 229·714 | 76·571 | 3 | 240·456 | 0·188 |
|  | ns(Day, df = 3):Priming | 0·024 | 3914·043 | 1304·681 | 3 | 240·754 | 3·208 |
|  | ns(Day, df = 3):Outcome | 0·798 | 1252·62 | 208·77 | 6 | 242·098 | 0·513 |

#### Table S3. Mixed-effect model of Serum IgG

| Antigen | Factor | P-value | Sum·Sq | Mean·Sq | DF | DenDF | F·value |
| --- | --- | --- | --- | --- | --- | --- | --- |
| PT | CFU_Scale | 0·709 | 4·121 | 4·121 | 1 | 169·076 | 0·14 |
|  | ns(Day, df = 3) | <0·001 | 915·445 | 305·148 | 3 | 371·914 | 10·375 |
|  | Priming | 0·980 | 0·019 | 0·019 | 1 | 170·55 | <0·001 |
|  | Outcome | 0·982 | 1·087 | 0·543 | 2 | 177·732 | 0·018 |
|  | CFU_Scale:ns(Day, df = 3) | 0·458 | 76·547 | 25·516 | 3 | 370·87 | 0·868 |
|  | ns(Day, df = 3):Priming | <0·001 | 535·251 | 178·417 | 3 | 372·23 | 6·066 |
|  | ns(Day, df = 3):Outcome | 0·001 | 667·271 | 111·212 | 6 | 371·443 | 3·781 |
| PRN | CFU_Scale | 0·835 | 2·369 | 2·369 | 1 | 177·293 | 0·043 |
|  | ns(Day, df = 3) | <0·001 | 909·22 | 303·073 | 3 | 372·539 | 5·535 |
|  | Priming | 0·788 | 3·966 | 3·966 | 1 | 178·763 | 0·072 |
|  | Outcome | 0·99 | 1·052 | 0·526 | 2 | 186·184 | 0·01 |
|  | CFU_Scale:ns(Day, df = 3) | 0·723 | 72·613 | 24·204 | 3 | 371·481 | 0·442 |
|  | ns(Day, df = 3):Priming | 0·247 | 227·738 | 75·913 | 3 | 372·891 | 1·386 |
|  | ns(Day, df = 3):Outcome | 0·711 | 205·238 | 34·206 | 6 | 372·056 | 0·625 |
| FHA | CFU_Scale | 0·973 | 0·015 | 0·015 | 1 | 126·323 | 0·001 |
|  | ns(Day, df = 3) | <0·001 | 307·718 | 102·573 | 3 | 369·672 | 7·701 |
|  | Priming | 0·982 | 0·007 | 0·007 | 1 | 127·569 | <0·001 |
|  | Outcome | 0·891 | 3·072 | 1·536 | 2 | 132·756 | 0·115 |
|  | CFU_Scale:ns(Day, df = 3) | 0·608 | 24·403 | 8·134 | 3 | 368·806 | 0·611 |
|  | ns(Day, df = 3):Priming | 0·985 | 2·034 | 0·678 | 3 | 369·785 | 0·051 |
|  | ns(Day, df = 3):Outcome | 0·059 | 163·154 | 27·192 | 6 | 369·31 | 2·042 |
| FIM | CFU_Scale | 0·89 | 0·185 | 0·185 | 1 | 171·895 | 0·019 |
|  | ns(Day, df = 3) | 0·001 | 158·131 | 52·71 | 3 | 372·737 | 5·498 |
|  | Priming | 0·796 | 0·641 | 0·641 | 1 | 173·374 | 0·067 |
|  | Outcome | 0·985 | 0·285 | 0·142 | 2 | 180·605 | 0·015 |
|  | CFU_Scale:ns(Day, df = 3) | 0·222 | 42·35 | 14·117 | 3 | 371·708 | 1·472 |
|  | ns(Day, df = 3):Priming | 0·317 | 33·954 | 11·318 | 3 | 373·053 | 1·181 |
|  | ns(Day, df = 3):Outcome | 0·119 | 97·983 | 16·331 | 6 | 372·273 | 1·703 |
| ACT | CFU_Scale | 0·918 | 0·181 | 0·181 | 1 | 145·387 | 0·011 |
|  | ns(Day, df = 3) | <0·001 | 860·295 | 286·765 | 3 | 369·367 | 16·867 |
|  | Priming | 0·98 | 0·011 | 0·011 | 1 | 146·794 | <0·001 |
|  | Outcome | 0·95 | 1·729 | 0·865 | 2 | 153·113 | 0·051 |
|  | CFU_Scale:ns(Day, df = 3) | 0·415 | 48·621 | 16·207 | 3 | 368·363 | 0·953 |
|  | ns(Day, df = 3):Priming | <0·001 | 761·985 | 253·995 | 3 | 369·592 | 14·94 |
|  | ns(Day, df = 3):Outcome | 0·07 | 199·959 | 33·327 | 6 | 368·93 | 1·96 |

#### Table S4. Mixed-effect model of Nasal IgA

| Antigen | Factor | Sum·Sq | Mean·Sq | NumDF | DenDF | F·value | Pr··F· |
| --- | --- | --- | --- | --- | --- | --- | --- |
| PT | CFU_Scale | 1·272 | 1·272 | 1 | 209·038 | 0·512 | 0·475 |
|  | ns(Day, df = 3) | 15·137 | 5·046 | 3 | 209·781 | 2·029 | 0·111 |
|  | Priming | 0·733 | 0·733 | 1 | 209·11 | 0·295 | 0·588 |
|  | Outcome | 2·5 | 1·25 | 2 | 209·193 | 0·503 | 0·606 |
|  | CFU_Scale:ns(Day, df = 3) | 70·792 | 23·597 | 3 | 210·913 | 9·492 | <0·001 |
|  | ns(Day, df = 3):Priming | 9·77 | 3·257 | 3 | 211·373 | 1·31 | 0·272 |
|  | ns(Day, df = 3):Outcome | 98·466 | 16·411 | 6 | 209·672 | 6·601 | <0·001 |
| PRN | CFU_Scale | 3·57 | 3·57 | 1 | 187·94 | 0·101 | 0·751 |
|  | ns(Day, df = 3) | 637·087 | 212·362 | 3 | 211·34 | 5·991 | <0·001 |
|  | Priming | 0·017 | 0·017 | 1 | 188·039 | <0·001 | 0·983 |
|  | Outcome | 8·745 | 4·373 | 2 | 188·133 | 0·123 | 0·884 |
|  | CFU_Scale:ns(Day, df = 3) | 336·447 | 112·149 | 3 | 212·136 | 3·164 | 0·025 |
|  | ns(Day, df = 3):Priming | 263·255 | 87·752 | 3 | 212·499 | 2·475 | 0·062 |
|  | ns(Day, df = 3):Outcome | 491·615 | 81·936 | 6 | 211·241 | 2·311 | 0·035 |
| FHA | CFU_Scale | 23·635 | 23·635 | 1 | 182·959 | 0·181 | 0·671 |
|  | ns(Day, df = 3) | 4983·341 | 1661·114 | 3 | 211·271 | 12·724 | <0·001 |
|  | Priming | 3·493 | 3·493 | 1 | 183·064 | 0·027 | 0·87 |
|  | Outcome | 22·526 | 11·263 | 2 | 183·159 | 0·086 | 0·917 |
|  | CFU_Scale:ns(Day, df = 3) | 991·11 | 330·37 | 3 | 212·022 | 2·531 | 0·058 |
|  | ns(Day, df = 3):Priming | 1350·247 | 450·082 | 3 | 212·371 | 3·448 | 0·018 |
|  | ns(Day, df = 3):Outcome | 2566·339 | 427·723 | 6 | 211·174 | 3·276 | 0·004 |
| FIM | CFU_Scale | 0·806 | 0·806 | 1 | 178·804 | 0·012 | 0·914 |
|  | ns(Day, df = 3) | 1252·022 | 417·341 | 3 | 211·275 | 6·059 | <0·001 |
|  | Priming | 1·028 | 1·028 | 1 | 178·912 | 0·015 | 0·903 |
|  | Outcome | 9·58 | 4·79 | 2 | 179·008 | 0·07 | 0·933 |
|  | CFU_Scale:ns(Day, df = 3) | 74·849 | 24·95 | 3 | 211·989 | 0·362 | 0·78 |
|  | ns(Day, df = 3):Priming | 231·625 | 77·208 | 3 | 212·326 | 1·121 | 0·342 |
|  | ns(Day, df = 3):Outcome | 810·347 | 135·058 | 6 | 211·179 | 1·961 | 0·073 |
| ACT | CFU_Scale | 11·714 | 11·714 | 1 | 201·741 | 0·304 | 0·582 |
|  | ns(Day, df = 3) | 722·078 | 240·693 | 3 | 204·505 | 6·251 | <0·001 |
|  | Priming | 0·919 | 0·919 | 1 | 202·057 | 0·024 | 0·877 |
|  | Outcome | 6·541 | 3·27 | 2 | 201·971 | 0·085 | 0·919 |
|  | CFU_Scale:ns(Day, df = 3) | 955·44 | 318·48 | 3 | 205·442 | 8·271 | <0·001 |
|  | ns(Day, df = 3):Priming | 131·578 | 43·859 | 3 | 205·913 | 1·139 | 0·334 |
|  | ns(Day, df = 3):Outcome | 654·12 | 109·02 | 6 | 204·268 | 2·831 | 0·011 |

Table S5. Mixed-effect model of Nasal IgG

| Antigen | Factor | Sum·Sq | Mean·Sq | NumDF | DenDF | F·value | Pr··F· |
| --- | --- | --- | --- | --- | --- | --- | --- |
| PT | CFU_Scale | 1·586 | 1·586 | 1 | 162·726 | 0·057 | 0·812 |
|  | ns(Day, df = 3) | 384·635 | 128·212 | 3 | 210·841 | 4·61 | 0·004 |
|  | Priming | 0·371 | 0·371 | 1 | 162·843 | 0·013 | 0·908 |
|  | Outcome | 1·838 | 0·919 | 2 | 162·938 | 0·033 | 0·968 |
|  | CFU_Scale:ns(Day, df = 3) | 459·327 | 153·109 | 3 | 211·443 | 5·505 | 0·001 |
|  | ns(Day, df = 3):Priming | 124·209 | 41·403 | 3 | 211·744 | 1·489 | 0·219 |
|  | ns(Day, df = 3):Outcome | 544·22 | 90·703 | 6 | 210·751 | 3·261 | 0·004 |
| PRN | CFU_Scale | 0·068 | 0·068 | 1 | 195·013 | 0·016 | 0·898 |
|  | ns(Day, df = 3) | 52·978 | 17·659 | 3 | 211·805 | 4·22 | 0·006 |
|  | Priming | 0·026 | 0·026 | 1 | 195·105 | 0·006 | 0·937 |
|  | Outcome | 0·08 | 0·04 | 2 | 195·195 | 0·01 | 0·99 |
|  | CFU_Scale:ns(Day, df = 3) | 22·224 | 7·408 | 3 | 212·657 | 1·77 | 0·154 |
|  | ns(Day, df = 3):Priming | 5·427 | 1·809 | 3 | 213·035 | 0·432 | 0·73 |
|  | ns(Day, df = 3):Outcome | 59·317 | 9·886 | 6 | 211·705 | 2·363 | 0·031 |
| FHA | CFU_Scale | 0·108 | 0·108 | 1 | 116·755 | 0·013 | 0·909 |
|  | ns(Day, df = 3) | 186·98 | 62·327 | 3 | 207·182 | 7·563 | <0·001 |
|  | Priming | 0·056 | 0·056 | 1 | 116·864 | 0·007 | 0·934 |
|  | Outcome | 0·885 | 0·442 | 2 | 116·94 | 0·054 | 0·948 |
|  | CFU_Scale:ns(Day, df = 3) | 4·744 | 1·581 | 3 | 207·594 | 0·192 | 0·902 |
|  | ns(Day, df = 3):Priming | 20·166 | 6·722 | 3 | 207·825 | 0·816 | 0·487 |
|  | ns(Day, df = 3):Outcome | 72·223 | 12·037 | 6 | 207·107 | 1·461 | 0·193 |
| FIM | CFU_Scale | 0·01 | 0·01 | 1 | 196·155 | 0·001 | 0·971 |
|  | ns(Day, df = 3) | 147·514 | 49·171 | 3 | 212·41 | 6·238 | <0·001 |
|  | Priming | 0·184 | 0·184 | 1 | 196·246 | 0·023 | 0·879 |
|  | Outcome | 0·273 | 0·136 | 2 | 196·335 | 0·017 | 0·983 |
|  | CFU_Scale:ns(Day, df = 3) | 1·121 | 0·374 | 3 | 213·249 | 0·047 | 0·986 |
|  | ns(Day, df = 3):Priming | 14·05 | 4·683 | 3 | 213·621 | 0·594 | 0·619 |
|  | ns(Day, df = 3):Outcome | 79·51 | 13·252 | 6 | 212·312 | 1·681 | 0·127 |
| ACT | CFU_Scale | 0·547 | 0·547 | 1 | 161·456 | 0·195 | 0·659 |
|  | ns(Day, df = 3) | 88·439 | 29·48 | 3 | 203·631 | 10·509 | <0·001 |
|  | Priming | 0·069 | 0·069 | 1 | 162·035 | 0·025 | 0·875 |
|  | Outcome | 0·236 | 0·118 | 2 | 161·842 | 0·042 | 0·959 |
|  | CFU_Scale:ns(Day, df = 3) | 38·401 | 12·8 | 3 | 204·275 | 4·563 | 0·004 |
|  | ns(Day, df = 3):Priming | 14·863 | 4·954 | 3 | 204·451 | 1·766 | 0·155 |
|  | ns(Day, df = 3):Outcome | 29·998 | 5 | 6 | 203·729 | 1·782 | 0·104 |

Table S6. AUC and thresholds distinguishing non-infected vs shedding

Bolded values represent factors that pass the significance thresholding [AUC≥0·70, Youden index≥0·30, sensitivity>0·70, and specificity>0·50]. AUC and Cutpoint reported as median (95% CI).

|  |  | AUC | Sensitivity | Specificity | Youden | Cutpoint |
| --- | --- | --- | --- | --- | --- | --- |
| Baseline (Day -1) | | | | | | |
| PT | Serum IgA | 0·607 (0·458-0·756) | 0·833 | 0·500 | 0·333 | 1·74 (1·74-1·74) |
|  | Serum IgG | 0·601 (0·422-0·78) | 0·667 | 0·319 | -0·014 | 3·4 (3·4-4·73) |
|  | Nasal Wash IgG | 0·77 (0·498-1) | 0·500 | 0·941 | 0·441 | 0·004 (0·004-0·004) |
|  | Nasal Wash IgA | 0·588 (0·381-0·795) | 0·667 | 0·559 | 0·225 | 0·002 (0·002-0·002) |
| PRN | Serum IgA | 0·707 (0·563-0·852) | 0·667 | 0·426 | 0·092 | 9·14 (2·61-9·14) |
|  | Serum IgG | 0·601 (0·396-0·806) | 0·583 | 0·723 | 0·307 | 78·89 (78·89-78·89) |
|  | Nasal Wash IgG | 0·672 (0·434-0·909) | 0·667 | 0·647 | 0·314 | 0·0132 (0·0132-0·0132) |
|  | Nasal Wash IgA | 0·738 (0·502-0·973) | 0·500 | 0·824 | 0·324 | 0·005 (0·005-0·005) |
| FHA | **Serum IgA** | **0·752 (0·622-0·882)** | **0·750** | **0·660** | **0·410** | **5·55 (5·55-5·55)** |
|  | Serum IgG | 0·606 (0·414-0·799) | 0·417 | 0·489 | -0·094 | 23·86 (23·86-42·29) |
|  | **Nasal Wash IgG** | **0·765 (0·604-0·926)** | **0·833** | **0·559** | **0·392** | **0·0056 (0·0056-0·0056)** |
|  | Nasal Wash IgA | 0·833 (0·683-0·983) | 0·667 | 0·618 | 0·284 | 0·0017 (0·0017-0·002) |
| FIM | **Serum IgA** | **0·764 (0·635-0·894)** | **0·750** | **0·574** | **0·324** | **5·28 (5·28-7·13)** |
|  | Serum IgG | 0·571 (0·38-0·762) | 0·500 | 0·766 | 0·266 | 41·29 (18·97-41·29) |
|  | Nasal Wash IgG | 0·397 (0·139-0·655) | 0·500 | 0·412 | -0·088 | 0·0035 (0·0034-0·0109) |
|  | Nasal Wash IgA | 0·728 (0·509-0·947) | 0·833 | 0·500 | 0·333 | 0·0016 (0·0016-0·0016) |
| ACT | Serum IgG | 0·468 (0·268-0·668) | 0·833 | 0·170 | 0·004 | 1228·35 (653·96-2366·88) |
|  | Nasal Wash IgG | 0·561 (0·265-0·856) | 0·500 | 0·788 | 0·288 | 3·78 (0·91-3·78) |
|  | Nasal Wash IgA | 0·51 (0·225-0·796) | 0·500 | 0·485 | -0·015 | 0·2 (0·199-0·2005) |
| Day 1 | | | | | | |
| PT | Serum IgG | 0·696 (0·489-0·904) | 0·857 | 0·375 | 0·232 | 3·95 (3·95-3·95) |
|  | Nasal Wash IgG | 0·629 (0·395-0·862) | 0·429 | 0·250 | -0·321 | 0·0014 (7e-04-0·0024) |
|  | Nasal Wash IgA | 0·605 (0·383-0·827) | 0·429 | 0·250 | -0·321 | 0·0015 (0·001-0·0021) |
| PRN | Serum IgG | 0·618 (0·357-0·878) | 0·571 | 0·700 | 0·271 | 73·57 (73·57-73·57) |
|  | Nasal Wash IgG | 0·661 (0·444-0·877) | 0·571 | 0·525 | 0·096 | 0·0083 (0·0083-0·0123) |
|  | **Nasal Wash IgA** | **0·737 (0·579-0·896)** | **0·857** | **0·550** | **0·407** | **0·0018 (0·0018-0·0018)** |
| FHA | Serum IgG | 0·814 (0·669-0·96) | 0·571 | 0·550 | 0·121 | 29·57 (29·57-49·09) |
|  | **Nasal Wash IgG** | **0·754 (0·587-0·92)** | **0·714** | **0·650** | **0·364** | **0·0071 (0·0071-0·0071)** |
|  | Nasal Wash IgA | 0·773 (0·585-0·961) | 0·429 | 0·500 | -0·071 | 8e-04 (8e-04-0·0029) |
| FIM | Serum IgG | 0·414 (0·169-0·659) | 0·429 | 0·300 | -0·271 | 13·82 (13·82-13·82) |
|  | Nasal Wash IgG | 0·343 (0·104-0·582) | 0·571 | 0·375 | -0·054 | 0·004 (0·0016-0·004) |
|  | Nasal Wash IgA | 0·677 (0·486-0·868) | 0·857 | 0·400 | 0·257 | 0·0015 (0·0015-0·0015) |
| ACT | Serum IgG | 0·654 (0·456-0·851) | 0·714 | 0·350 | 0·064 | 2226·4 (2226·4-3052·12) |
|  | Nasal Wash IgG | 0·626 (0·404-0·848) | 0·429 | 0·410 | -0·161 | 3·49 (0·73-3·49) |
|  | Nasal Wash IgA | 0·549 (0·266-0·833) | 0·143 | 0·974 | 0·117 | 0·59 (0·17-0·59) |
| Day 3 | | | | | | |
| PT | Serum IgA | 0·528 (0·359-0·697) | 0·286 | 0·500 | -0·214 | 3·09 (1·52-3·75) |
|  | **Serum IgG** | **0·744 (0·56-0·929)** | **0·857** | **0·526** | **0·383** | **4·81 (4·81-4·81)** |
|  | Nasal Wash IgG | 0·696 (0·483-0·909) | 0·667 | 0·575 | 0·242 | 0·0016 (0·0016-0·0016) |
|  | Nasal Wash IgA | 0·738 (0·464-1) | 0·500 | 0·850 | 0·350 | 0·0027 (0·0027-0·0027) |
| PRN | Serum IgA | 0·737 (0·553-0·922) | 0·571 | 0·730 | 0·301 | 8·93 (8·93-8·93) |
|  | Serum IgG | 0·643 (0·384-0·901) | 0·571 | 0·737 | 0·308 | 78·61 (78·61-78·61) |
|  | Nasal Wash IgG | 0·621 (0·372-0·87) | 0·333 | 0·600 | -0·067 | 0·0224 (0·0086-0·0224) |
|  | Nasal Wash IgA | 0·719 (0·51-0·927) | 0·500 | 0·575 | 0·075 | 0·0022 (0·0022-0·004) |
| FHA | Serum IgA | 0·792 (0·632-0·951) | 0·571 | 0·838 | 0·409 | 15·96 (15·96-15·96) |
|  | **Serum IgG** | **0·82 (0·696-0·943)** | **0·857** | **0·684** | **0·541** | **34·37 (34·37-34·37)** |
|  | **Nasal Wash IgG** | **0·779 (0·637-0·921)** | **0·833** | **0·625** | **0·458** | **0·006 (0·006-0·006)** |
|  | Nasal Wash IgA | 0·812 (0·612-1) | 0·500 | 0·725 | 0·225 | 0·0019 (0·0019-0·0034) |
| FIM | **Serum IgA** | **0·734 (0·579-0·888)** | **0·714** | **0·676** | **0·390** | **6·15 (6·15-6·15)** |
|  | Serum IgG | 0·417 (0·154-0·68) | 0·429 | 0·316 | -0·256 | 16·04 (16·04-16·04) |
|  | Nasal Wash IgG | 0·321 (0·002-0·639) | 0·500 | 0·200 | -0·300 | 0·0018 (0·0018-0·0018) |
|  | Nasal Wash IgA | 0·652 (0·449-0·855) | 0·667 | 0·375 | 0·042 | 0·0016 (0·0016-0·002) |
| ACT | Serum IgG | 0·647 (0·457-0·836) | 0·857 | 0·395 | 0·252 | 2295·59 (2295·59-2295·59) |
|  | Nasal Wash IgG | 0·621 (0·345-0·896) | 0·500 | 0·875 | 0·375 | 3·81 (0·91-3·81) |
|  | Nasal Wash IgA | 0·548 (0·227-0·869) | 0·500 | 0·975 | 0·475 | 0·62 (0·17-0·62) |
| Day 7 | | | | | | |
| PT | Serum IgA | 0·546 (0·389-0·702) | 0·857 | 0·487 | 0·344 | 1·69 (1·69-1·69) |
|  | Serum IgG | 0·7 (0·486-0·914) | 0·429 | 0·350 | -0·221 | 3·95 (3·95-14·94) |
|  | Nasal Wash IgG | 0·661 (0·441-0·881) | 0·857 | 0·400 | 0·257 | 7e-04 (7e-04-7e-04) |
|  | Nasal Wash IgA | 0·633 (0·387-0·878) | 0·286 | 0·743 | 0·029 | 0·0024 (0·0024-0·0072) |
| PRN | Serum IgA | 0·733 (0·564-0·901) | 0·571 | 0·795 | 0·366 | 9·92 (9·92-9·92) |
|  | Serum IgG | 0·643 (0·389-0·897) | 0·571 | 0·700 | 0·271 | 75·47 (75·47-75·47) |
|  | Nasal Wash IgG | 0·592 (0·338-0·846) | 0·286 | 0·543 | -0·171 | 0·0125 (0·0091-0·0301) |
|  | Nasal Wash IgA | 0·692 (0·465-0·918) | 0·429 | 0·943 | 0·371 | 0·0061 (0·0016-0·0061) |
| FHA | Serum IgA | 0·791 (0·634-0·948) | 0·571 | 0·821 | 0·392 | 15·15 (9·44-15·15) |
|  | Serum IgG | 0·807 (0·67-0·944) | 0·714 | 0·575 | 0·289 | 31·56 (31·56-34·75) |
|  | **Nasal Wash IgG** | **0·767 (0·619-0·916)** | **0·857** | **0·629** | **0·486** | **0·0057 (0·0057-0·0057)** |
|  | Nasal Wash IgA | 0·739 (0·472-1) | 0·571 | 0·829 | 0·400 | 0·0024 (0·0024-0·0024) |
| FIM | **Serum IgA** | **0·722 (0·554-0·89)** | **0·714** | **0·615** | **0·330** | **6·36 (6·36-6·36)** |
|  | Serum IgG | 0·393 (0·14-0·646) | 0·429 | 0·275 | -0·296 | 14·59 (14·59-14·59) |
|  | Nasal Wash IgG | 0·412 (0·173-0·651) | 0·429 | 0·371 | -0·200 | 0·0038 (0·0038-0·0038) |
|  | Nasal Wash IgA | 0·7 (0·523-0·877) | 0·857 | 0·486 | 0·343 | 0·0019 (0·0019-0·0019) |
| ACT | Serum IgG | 0·643 (0·444-0·842) | 0·857 | 0·375 | 0·232 | 2288·99 (2288·99-2288·99) |
|  | Nasal Wash IgG | 0·637 (0·421-0·852) | 0·571 | 0·686 | 0·257 | 1·63 (1·63-1·63) |
|  | Nasal Wash IgA | 0·514 (0·24-0·789) | 0·429 | 0·486 | -0·086 | 0·15 (0·15-0·67) |
| Day 14 | | | | | | |
| PT | Serum IgA | 0·555 (0·397-0·714) | 0·818 | 0·304 | 0·123 | 1·29 (1·29-1·57) |
|  | Serum IgG | 0·586 (0·387-0·785) | 0·455 | 0·255 | -0·290 | 4·32 (1·39-8·75) |
|  | Nasal Wash IgG | 0·784 (0·478-1) | 0·667 | 0·784 | 0·450 | 0·0021 (0·0021-0·0021) |
|  | Nasal Wash IgA | 0·651 (0·348-0·953) | 0·333 | 0·919 | 0·252 | 0·0048 (0·0048-0·0048) |
| PRN | Serum IgA | 0·708 (0·547-0·868) | 0·636 | 0·739 | 0·375 | 8·49 (8·49-8·49) |
|  | Serum IgG | 0·623 (0·421-0·825) | 0·455 | 0·723 | 0·178 | 82·43 (82·43-82·7) |
|  | Nasal Wash IgG | 0·712 (0·458-0·965) | 0·667 | 0·676 | 0·342 | 0·0117 (0·0117-0·0117) |
|  | Nasal Wash IgA | 0·685 (0·468-0·902) | 0·833 | 0·459 | 0·293 | 0·002 (0·002-0·002) |
| FHA | Serum IgA | 0·69 (0·534-0·846) | 0·636 | 0·348 | -0·016 | 5·46 (3·65-8·67) |
|  | Serum IgG | 0·576 (0·373-0·779) | 0·455 | 0·766 | 0·221 | 55·81 (29·37-55·81) |
|  | Nasal Wash IgG | 0·68 (0·463-0·898) | 0·500 | 0·378 | -0·122 | 0·0055 (0·0055-0·0096) |
|  | Nasal Wash IgA | 0·644 (0·331-0·958) | 0·333 | 0·946 | 0·279 | 0·0129 (0·0129-0·0129) |
| FIM | **Serum IgA** | **0·759 (0·619-0·899)** | **0·818** | **0·674** | **0·492** | **9·44 (9·44-9·44)** |
|  | Serum IgG | 0·582 (0·375-0·79) | 0·455 | 0·787 | 0·242 | 46·95 (46·95-46·95) |
|  | Nasal Wash IgG | 0·356 (0·092-0·619) | 0·500 | 0·297 | -0·203 | 0·0013 (0·0013-0·003) |
|  | **Nasal Wash IgA** | **0·723 (0·557-0·889)** | **0·833** | **0·622** | **0·455** | **0·0033 (0·0033-0·0033)** |
| ACT | Serum IgG | 0·445 (0·232-0·658) | 0·727 | 0·170 | -0·103 | 902·37 (902·37-1964·95) |
|  | Nasal Wash IgG | 0·606 (0·283-0·93) | 0·500 | 0·889 | 0·389 | 4·92 (0·78-4·92) |
|  | Nasal Wash IgA | 0·48 (0·151-0·808) | 0·833 | 0·351 | 0·185 | 0·15 (0·131-0·15) |
| Day 28 | | | | | | |
| PT | Serum IgA | 0·394 (0·237-0·552) | 0·222 | 0·650 | -0·128 | 4·12 (4·12-5·88) |
|  | Serum IgG | 0·331 (0·145-0·516) | 0·444 | 0·425 | -0·131 | 9·59 (7·68-9·59) |
|  | Nasal Wash IgG | 0·401 (0·186-0·615) | 0·286 | 0·722 | 0·008 | 0·0074 (0·0048-0·0074) |
|  | Nasal Wash IgA | 0·361 (0·118-0·605) | 0·714 | 0·444 | 0·159 | 0·0023 (0·0012-0·0052) |
| PRN | Serum IgA | 0·433 (0·243-0·623) | 0·333 | 0·775 | 0·108 | 37·05 (9·21-37·05) |
|  | Serum IgG | 0·383 (0·146-0·621) | 0·556 | 0·100 | -0·344 | 22·48 (22·48-76·5) |
|  | Nasal Wash IgG | 0·504 (0·247-0·761) | 0·429 | 0·583 | 0·012 | 0·0221 (0·012-0·071) |
|  | Nasal Wash IgA | 0·353 (0·156-0·551) | 0·429 | 0·694 | 0·123 | 0·0145 (0·0028-0·0145) |
| FHA | Serum IgA | 0·428 (0·208-0·647) | 0·667 | 0·450 | 0·117 | 5·66 (5·66-16·17) |
|  | Serum IgG | 0·322 (0·127-0·517) | 0·444 | 0·700 | 0·144 | 96·59 (38·99-96·59) |
|  | Nasal Wash IgG | 0·369 (0·154-0·584) | 0·286 | 0·361 | -0·353 | 0·0101 (0·0101-0·0101) |
|  | Nasal Wash IgA | 0·278 (0·072-0·484) | 0·286 | 0·417 | -0·298 | 0·0063 (0·0063-0·0063) |
| FIM | Serum IgA | 0·567 (0·389-0·744) | 0·667 | 0·900 | 0·567 | 49·66 (5·36-81·03) |
|  | Serum IgG | 0·394 (0·183-0·606) | 0·556 | 0·275 | -0·169 | 19·52 (16·01-26·15) |
|  | Nasal Wash IgG | 0·377 (0·135-0·619) | 0·571 | 0·583 | 0·155 | 0·0106 (0·0045-0·0106) |
|  | Nasal Wash IgA | 0·387 (0·191-0·583) | 0·143 | 0·444 | -0·413 | 0·0068 (0·0068-0·0068) |
| ACT | Serum IgG | 0·231 (0·062-0·399) | 0·444 | 0·425 | -0·131 | 6629·1 (3992·72-6629·1) |
|  | Nasal Wash IgG | 0·504 (0·233-0·775) | 0·429 | 0·306 | -0·266 | 1·81 (1·81-5·12) |
|  | Nasal Wash IgA | 0·26 (0·077-0·443) | 0·286 | 0·306 | -0·409 | 0·25 (0·25-0·69) |
| Day 56 | | | | | | |
| PT | Serum IgA | 0·291 (0·114-0·467) | 0·375 | 0·459 | -0·166 | 3·75 (2·97-3·75) |
|  | Serum IgG | 0·226 (0·049-0·404) | 0·250 | 0·324 | -0·426 | 8·59 (8·59-8·59) |
|  | Nasal Wash IgG | 0·258 (0·093-0·423) | 0·286 | 0·528 | -0·187 | 0·0052 (0·0036-0·0068) |
|  | Nasal Wash IgA | 0·175 (0-0·409) | 0·286 | 0·111 | -0·603 | 0·0015 (0·0015-0·0018) |
| PRN | Serum IgA | 0·429 (0·223-0·636) | 0·250 | 0·757 | 0·007 | 37·42 (9·59-37·42) |
|  | Serum IgG | 0·345 (0·126-0·563) | 0·625 | 0·081 | -0·294 | 18·28 (18·28-87·19) |
|  | Nasal Wash IgG | 0·345 (0·131-0·56) | 0·286 | 0·444 | -0·270 | 0·0145 (0·0145-0·0145) |
|  | Nasal Wash IgA | 0·274 (0·087-0·461) | 0·429 | 0·278 | -0·294 | 0·0029 (0·0029-0·0029) |
| FHA | Serum IgA | 0·334 (0·152-0·517) | 0·250 | 0·649 | -0·101 | 25·58 (19·12-25·58) |
|  | Serum IgG | 0·26 (0·085-0·435) | 0·125 | 0·541 | -0·334 | 82·61 (82·61-82·61) |
|  | Nasal Wash IgG | 0·202 (0·036-0·368) | 0·286 | 0·500 | -0·214 | 0·0145 (0·0098-0·0145) |
|  | Nasal Wash IgA | 0·202 (0·016-0·389) | 0·429 | 0·222 | -0·349 | 0·0037 (0·0037-0·0048) |
| FIM | Serum IgA | 0·507 (0·323-0·691) | 0·500 | 0·514 | 0·014 | 17·73 (9·41-54·29) |
|  | Serum IgG | 0·392 (0·176-0·608) | 0·625 | 0·270 | -0·105 | 21·59 (21·59-84·31) |
|  | Nasal Wash IgG | 0·31 (0·072-0·547) | 0·571 | 0·500 | 0·071 | 0·0064 (0·0028-0·0099) |
|  | Nasal Wash IgA | 0·29 (0·089-0·49) | 0·286 | 0·417 | -0·298 | 0·0061 (0·0048-0·0061) |
| ACT | Serum IgG | 0·23 (0·049-0·41) | 0·375 | 0·243 | -0·382 | 5118·41 (5118·41-5118·41) |
|  | Nasal Wash IgG | 0·397 (0·169-0·624) | 0·571 | 0·472 | 0·044 | 2·02 (2·02-4·88) |
|  | Nasal Wash IgA | 0·119 (0-0·251) | 0·429 | 0·417 | -0·155 | 0·3 (0·2145-0·475) |

### 3 Supplementary Figures

Figure S1. Pearson Correlation between serum and nasal IgA and IgG across time·

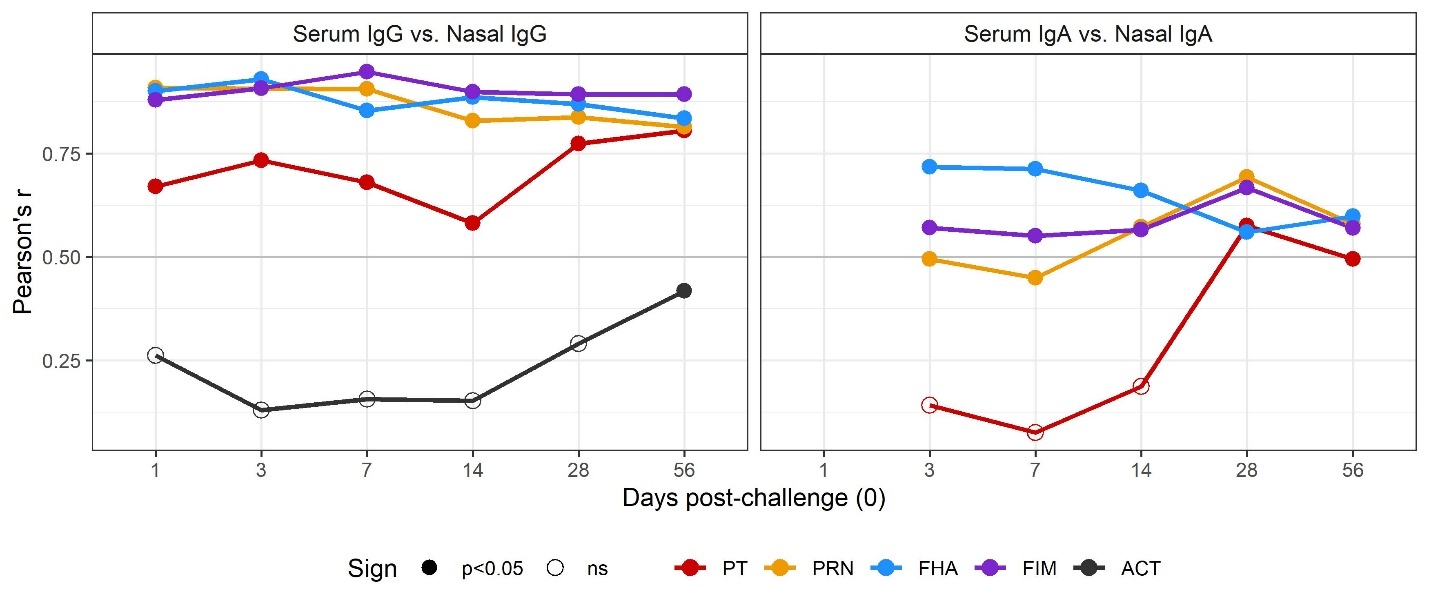
